# Convergent mutational processes in esophageal and junctional adenocarcinomas across eight countries

**DOI:** 10.64898/2026.10.21.26365720

**Authors:** Laura Torrens, Sarah Moody, Jiali Pang, Behnoush Abedi-Ardekani, Haoran Zhang, Ammal Abbasi, Charles Nunn, Stephen Fitzgerald, Pilar Gallego-García, Valerie Gaborieau, Thomas Cattiaux, Priscilia Chopard, Saamin Cheema, Emily Thomas, Calli Latimer, Christine Carreira, Marcos Díaz-Gay, Maria Paula Curado, Charles Dzamalala, Felipe Fermandes Coimbra, Lorenzo Ferri, Rebecca C Fitzgerald, Alisa M Goldstein, Antonio Huertas-Salgado, Reza Malekzadeh, Diana Menya, Rui Manuel Reis, Felipe Ribeiro Pinto, Gholamreza Roshandel, Jonathan Yeung, Ana Carolina de Carvalho, Laura Humphreys, Ayesha Noorani, Ludmil B Alexandrov, Paul Brennan, Michael R Stratton, Sandra Perdomo

**Author notes:** **CORRESPONDING AUTHOR** Correspondence to Sandra Perdomo and Michael R. Stratton. These authors contributed equally (Laura Torrens, Sarah Moody).

## Abstract

The incidence distribution of esophageal adenocarcinoma (EAC) and gastroesophageal junction (GEJ) cancers presents important geographic variations, yet the causes of these differences remain unclear. We analyzed 520 whole genomes from EAC/GEJ tumors across eight countries to examine the mutational forces shaping these cancers and their geographic variation. The mutational landscape was highly similar between EAC and GEJ tumors, and was dominated by signatures SBS17a/b, previously associated with reflux in EAC/GEJ. SBS17a/b were linked to obesity in EAC, supporting an indirect role in exacerbating reflux-mediated mutagenesis. Other established risk factors, such as tobacco smoking, did not yield a direct mutational footprint, pointing to non-mutagenic effects in EAC/GEJ. Nonetheless, geographic differences were present, with signatures related to endogenous cellular functions enriched in high-incidence populations. These findings provide new insights into the shared mutational forces that shape EAC and GEJ, as well as their geographic distribution.

## INTRODUCTION

Adenocarcinomas of the esophagus and gastroesophageal junction are common malignancies worldwide, and a leading cause of mortality, with five-year survival rates below 20%^1–3^. Globally, the annual incidence is estimated at 85,700 cases for esophageal adenocarcinoma (EAC) and 181,000 for gastroesophageal junction cancer (GEJ)^4,5^, although both exhibit important variations in their geographic distribution. Regions in central China report the world’s highest incidence of GEJ cancer, reaching rates up to fivefold above the national average, whereas North America and Western Europe, and in particular the United Kingdom, carry the highest EAC burdens^4,6^. The underlying causes of this distribution remain unclear, although differences in the prevalence of gastroesophageal reflux disease (GERD) and obesity have been proposed as major contributors^4,5^. Despite this global variation, large-scale genomic studies have been mostly restricted to single populations within Europe, North America, or China^7–11^, hindering our ability to map the distribution of mutagenic processes and the potential impact on cancer risk. To address this, the Mutographs Cancer Grand Challenge project^12^ leverages a strategy that integrates mutational signature analysis with extensive epidemiological information across regions with varying incidence rates. This framework has been instrumental in defining the mutational landscapes in multiple other cancer types and identifying or ruling out the mutagenic impact of suspected carcinogens^13–16^.

The analysis of mutational signatures is an effective tool to track the exposure to endogenous and exogenous mutagenic factors over a patient’s lifetime. EAC and GEJ share multiple risk factors^4^, including GERD^17,18^, obesity^19,20^, and tobacco smoking^17,20^ Alcohol drinking has been suspected as an EAC/GEJ-related exposure, although the evidence is not conclusive^21,22^. Previous studies have investigated the mutational signature landscape in EAC or GEJ^7,10,11^, but genomic epidemiology evidence linking specific mutational patterns to risk factors remains limited. In particular, SBS17 signatures (SBS17a and SBS17b), a hallmark feature of EAC/GEJ and precursor lesions, have been proposed to reflect mutagenesis arising from oxidized guanine nucleotides^23,24^. Although several oxidative mechanisms may be able to induce SBS17, gastroesophageal reflux is considered the leading candidate in EAC/GEJ^4,5^. However, a causal link between this exposure and SBS17 signatures has yet to be established^23–25^. Dissecting the mutagenic contributions of EAC/GEJ-related exposures remains an unmet challenge, leaving a significant gap in our understanding of how lifestyle factors translate into somatic alterations.

Despite their overlapping risk factors and anatomical proximity, clinical algorithms classify EAC and GEJ as esophageal and gastric tumors, respectively, according to their anatomical relationship to the gastroesophageal junction^26–28^. Specifically, EAC predominantly arises in the distal esophagus, whereas GEJ develops at the intersection of the esophagus and the proximal stomach, and is further subdivided into three groups under the Siewert classification^29^. EAC and Siewert type I GEJ arise from Barrett’s esophagus (BE) secondary to gastro-esophageal reflux, a metaplastic state in which the stratified squamous esophageal epithelium is replaced by a columnar, intestinal-type mucosa^30,31^. Conversely, Siewert type III tumors rarely exhibit a clear association with BE, whereas type II tumors display a mixed etiology^32^. Notably, the appropriate demarcation between gastric and esophageal adenocarcinomas remains unresolved, yet this distinction is critical for guiding prevention strategies and treatment decisions^26–28^. Determining the extent to which EAC and Siewert type I-III tumors share molecular underpinnings could therefore inform this classification and improve our understanding of how carcinogenic exposures drive carcinogenesis across anatomical subsites^28,29^.

Here, we present a comprehensive whole-genome sequencing (WGS) analysis of 520 EAC and GEJ tumors from eight countries across four continents. By integrating genomic, demographic, and exposure information, we characterize the mutational processes shaping these cancers and their potential implications in the global distribution of EAC/GEJ cancer incidence.

## RESULTS

### A transcontinental cohort of EAC and GEJ

We assembled a cohort of 520 EAC/GEJ cases from eight countries across diverse geographical regions and ancestry backgrounds from Europe (the United Kingdom), the Americas (Canada, Brazil, Colombia), Africa (Kenya, Malawi), and Asia (China, Iran) (**Figure 1a, Extended Data Figure 1a-c**). This cohort captures a wide range of disease incidence, including the high-incidence region in Shanxi, China (for GEJ) and the United Kingdom (for EAC) (**Figure 1b**). A total of 159 EAC and 361 GEJ cases were included in the cohort. GEJ cases were further classified according to the Siewert system (type I, n = 55, 15.2%; type II, n = 72, 19.9%; type III, n = 196, 54.3%; and unspecified, n = 38, 10.5%) (**Figure 1c**). Harmonized clinical and epidemiological data were collected for all participants, including diagnosis of GERD, obesity (defined as body mass index above 30), tobacco smoking, and alcohol consumption (**Figure 1d**).

**Figure 1.**
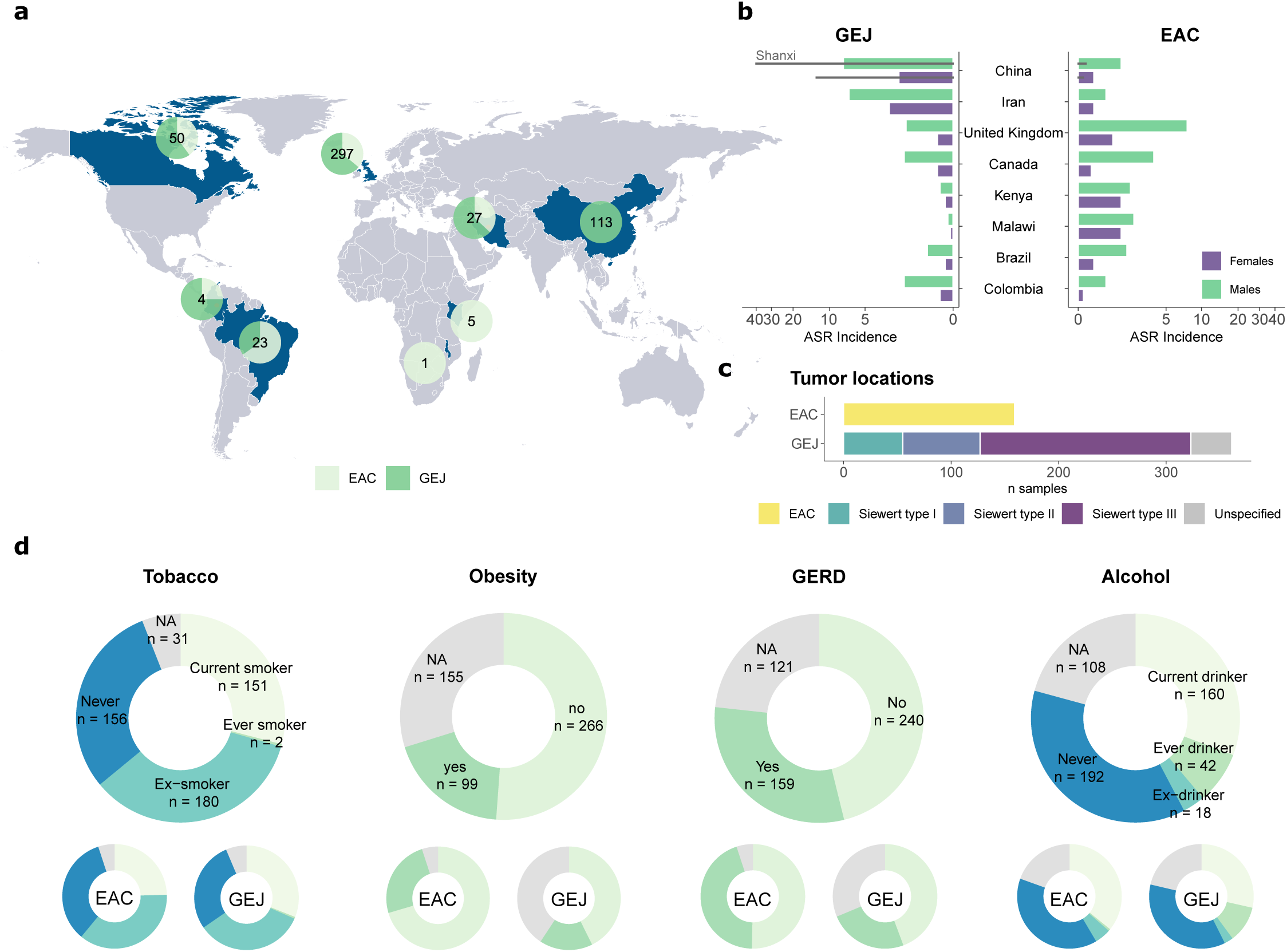
Geographic and epidemiological characterization of the Mutographs EAC/GEJ cohort. **a,** Map showing countries included in the study. Circles indicate the total number of cases per country and the proportion of EAC and GEJ cases. **b,** Incidence rates of EAC and GEJ across countries in males and females. Values for the high-incidence region in Shanxi, China, are shown in black. **c,** Sample distribution based on tumor location. **d,** Distribution of known or suspected risk factors based on epidemiological questionnaire data. Frequencies in the complete dataset (top) and by tumor subtype (bottom) are indicated.

Paired tumor and normal tissue DNA underwent whole-genome sequencing at a mean coverage of 56-fold and 27-fold, respectively. Of the 520 EAC/GEJ cases, 486 (93.5%) were categorized as microsatellite stable (MSS) based on their somatic mutation burdens and profiles. Microsatellite unstable (MSI) cases (n=23, 4.4%) and samples with deficiencies in other DNA repair pathways (10, 1.9%, including germline mutations in *BRCA1* (n=9) and *MUTYH* (n=1)) were excluded from further analyses to avoid interference with signature extraction^13,33^. One case with prior chemotherapy exposure was also discarded due to high treatment-related mutational burden^7^. The final analytical cohort comprised 486 treatment-naive, DNA repair-proficient EAC/GEJ (n = 151 EAC, n = 335 GEJ).

### Mutation burden profile in EAC and GEJ

In our cohort of 486 DNA repair-proficient EAC/GEJ, we observed a median of 17,536 single-base substitutions (SBSs; range = 2,897–171,939), 50 doublet-base substitutions (DBSs; range = 6–357), and 1,703 small insertions and deletions (IDs; range = 84-7,263) per genome (**Supplementary Table 1**). Small but significant differences were observed across geographical regions after correction for tumor purity and other relevant covariates, including higher ID burdens in China compared to other regions (**Extended Data Figure 2**, **Supplementary Table 2**). SBS, DBS, and ID burdens were overall homogeneous across diverse backgrounds, with no significant association between mutational burdens and exposure to known EAC/GEJ risk factors, such as tobacco smoking or obesity. Mutational loads were also comparable across anatomical sites, with the exception of significantly decreased ID burden in EAC relative to Siewert type II and III GEJ (**Extended Data Figure 2**, **Supplementary Table 2**).

### Mutational signature landscape of EAC/GEJ reveals dysregulations of endogenous cellular functions

To define the mutagenic forces driving EAC/GEJ development, we extracted SBS, DBS, and ID signatures and estimated the contribution of each signature to every sample. We obtained 12 *de novo* SBS signatures, which were decomposed into 13 reference signatures from the Catalog of Somatic Mutations in Cancer (COSMIC v3.6) database, and one signature previously described in colorectal cancer and pan-cancer datasets^13,34^, namely SBS111 (**Figure 2, Extended Data Figure 3, Supplementary Table 1Supplementary Table *3*, Supplementary Table *6*-Supplementary Table *8***, **Supplementary Note**). Signatures SBS17a and SBS17b accounted for 26.9% of the median SBS signature burden and were present in 97.6% of cases, reinforcing previous findings describing SBS17 signatures as the main genomic hallmark of these malignancies and in BE^7,10,11^. Signature SBS111, a signature with unknown etiology, which has been speculated to arise from DNA repair infidelity^34^, emerged as the second most abundant signature (median relative burden of 12%, found in 90% of cases), suggesting that this process is important for EAC/GEJ carcinogenesis. SBS102 and SBS103, two additional signatures of unknown etiology which have not been previously identified in EAC/GEJ^13^, were active in 71.0% and 12.8% of cases and contributed 8.2% and 0.9% to the median relative burden, respectively. Other common signatures included clock-like signatures SBS5 and SBS1, signatures of unknown etiology SBS8 and SBS93, previously found in stomach cancer^35^, and the reactive oxygen species (ROS)-related signature SBS18.

**Figure 2.**
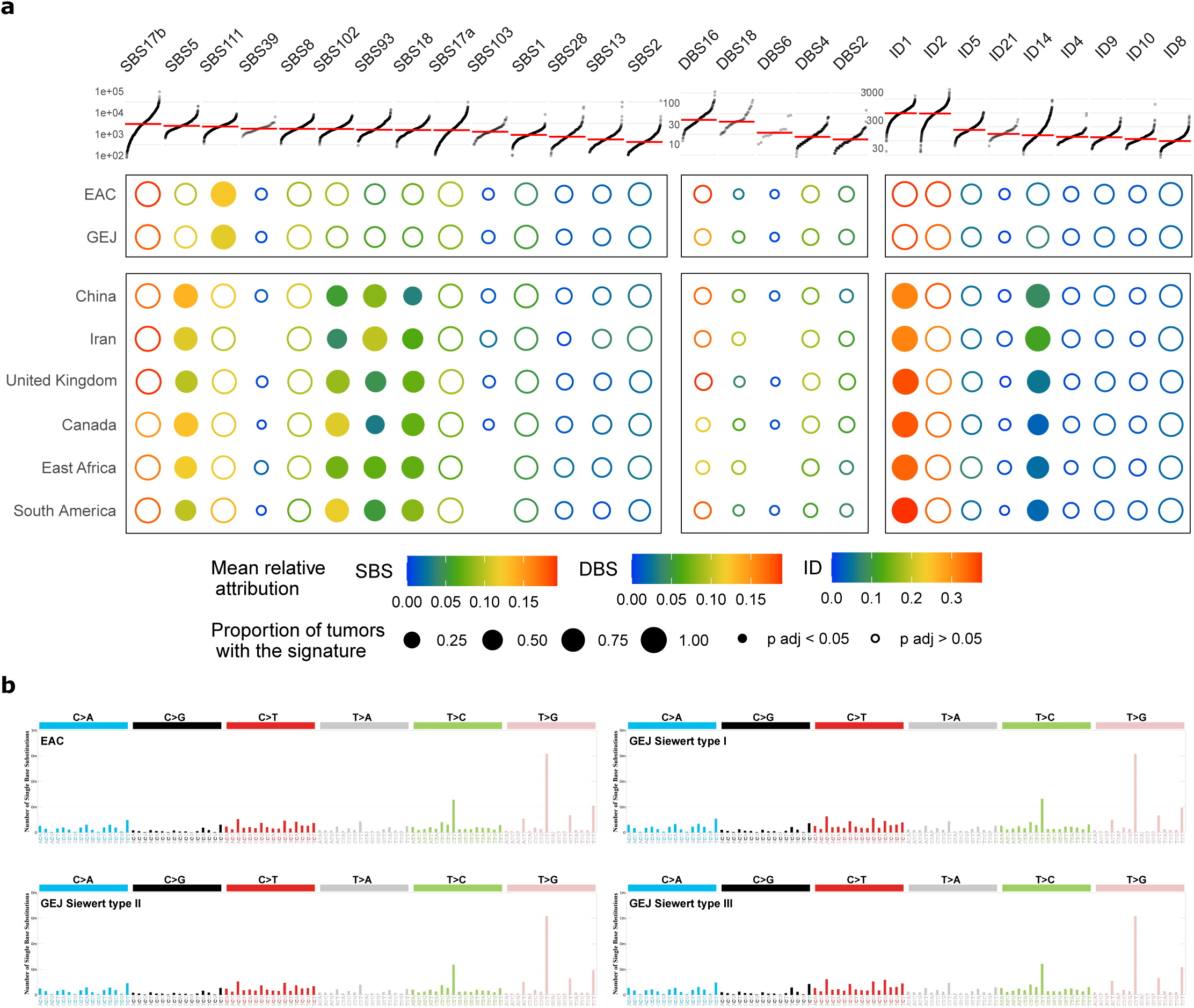
Mutational signature landscape of EAC/GEJ. **a**, SBS, DBS, and indel signatures extracted from 486 EAC/GEJ. The size of each dot represents the proportion of samples presenting each mutational signature in the whole dataset and across regions. The color represents the mean relative attribution of each signature. Dots filled in white indicate signatures without significantly different relative burdens across subsites. Significance was assessed using a two-sided Kruskal–Wallis test and Bonferroni correction, based on the total number of signatures considered per variant type. Top, the mutations per megabase attributed to each signature in samples with counts higher than zero. **b**, Mutational spectrum of EAC and GEJ.

Extraction of DBS and ID signatures identified three and five *de novo* signatures, respectively, which decomposed into five and nine COSMIC signatures (**Figure 2a**, **Extended Data Figure 4, Supplementary Table 4-Supplementary Table *8***, **Supplementary Note**). This included signatures DBS16 and ID5, both of which correlated with signatures SBS17a-b (**Extended Data Figure 4c**). Signature DBS16 has been previously identified in the esophagus and stomach and has been suggested to be related to SBS17 signatures^33^. Other relevant signature correlations included ID14 with SBS93, as observed in other tumor types^36^, and SBS111 with ID1 and ID2, related to replication slippage of the newly synthesized strand^7,34^.

Overall, the signature landscape of EAC/GEJ was dominated by oxidation damage, putatively secondary to reflux, and dysregulation of endogenous cellular functions. Despite the substantial mutation burden observed across the dataset and the wide variety of signatures identified, we found no evidence of signatures directly related to exogenous risk factors. Most notably, tobacco-specific signatures were absent, even though tobacco smoking was reported by 68% of patients in the cohort (333/489, **Figure 1d**).

### Timing of mutational processes during EAC/GEJ evolution

To reconstruct the temporal activity of mutagenic processes during EAC/GEJ development, we estimated the molecular timing of SBS and ID signatures (**Extended Data Figure 5a-b, Supplementary Table 9**). Acid reflux (SBS17a and b), clock-like (SBS1, SBS5), and ROS (SBS18) signatures were significantly enriched among early clonal mutations, consistent with mutagenic exposure occurring in pre-cancerous cells^37^, and in agreement with previous findings in EAC and in BE^11,37^. In contrast, several signatures were enriched in late clonal mutations, indicating increased activity during later tumor evolution stages. These included SBS103, APOBEC signatures, and signatures of unknown etiology (SBS39, SBS102, SBS8, SBS93, and ID4). Signature SBS111 was present both in early and late clonal mutations, indicating a sustained influence throughout the evolutionary trajectory of EAC/GEJ (**Extended Data Figure 5c**), as opposed to colorectal cancer, where it is predominantly a late event^34^. Importantly, EAC and GEJ displayed highly concordant evolutionary profiles (**Extended Data Figure 5d-e**).

### Signature differences across anatomical sites within the esophagus and junction

EAC and GEJ exhibited a remarkably similar mutational landscape and mutation burdens, with overlapping mutational profiles (**Figure 2b**, **Supplementary Table 2**), aligning with the shared epidemiological risk factors in both cancer subtypes. Consistent with this, principal component analysis of mutation counts showed no separation by tumor subtype (**Supplementary Note**). However, regression analysis revealed significant differences between EAC and GEJ in SBS111, as well as correlated ID signatures ID1 and ID2 (**Figure 3a,c**). While these signatures of DNA repair infidelity were present in both tumor subtypes, they were significantly enhanced in EAC tumors compared to GEJ.

**Figure 3.**
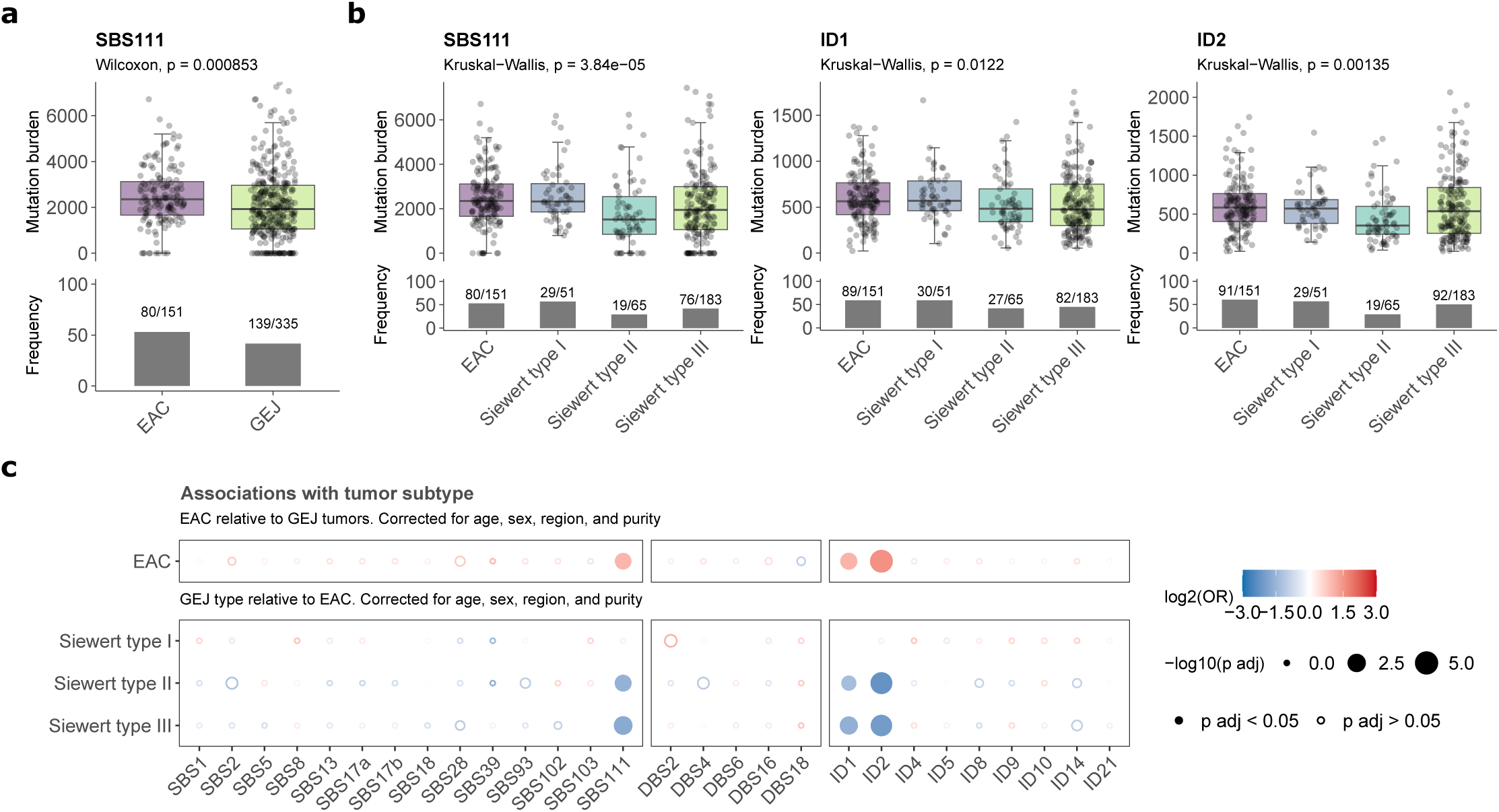
Differences in mutational signature profile across EAC and GEJ. **a-b,** Mutational burdens for signatures showing positive associations with EAC and GEJ tumor site (**a**) or across EAC and GEJ Siewert types (**b**) (n = 486 biologically independent samples). The Kruskal–Wallis test (two-sided) was used to test for global differences. Box-and-whisker plots are in the style of Tukey. The line within the box is plotted at the median, while the upper and lower ends indicate the 25th and 75th percentiles. Whiskers show 1.5× IQR. The y axes were cut at 1.25× upper whisker for clarity. Bar plots indicate the frequencies of dichotomized signatures. **c**, Logistic regression analyses measuring associations between mutational signatures and tumor subtypes. Regressions use either GEJ (top) or EAC (bottom) as reference. Dots filled in color indicate significant associations (adjusted p-value < 0.05). All regressions were corrected for age of diagnosis, sex, geographical region, and tumor purity. The Bonferroni method was used to adjust P values for multiple hypothesis testing, based on the total number of signatures considered per variant type.

To further refine this observation, we assessed the distribution of signatures across anatomical subsites using the Siewert classification. We observed an enrichment of SBS111, ID1, and ID2 in EAC and Siewert type I cases, corresponding to the lower and distal esophagus, relative to GEJ Siewert II and III tumors (**Figure 3b**). These associations were further confirmed by multivariate logistic regression analysis (**Figure 3c**, **Supplementary Table 10**), suggesting that, despite the overall mutational landscape of EAC and GEJ being highly similar, specific mutational patterns are more abundant in EAC and Siewert I GEJ compared to other GEJ subsites. The analysis of somatic driver and copy number alterations also revealed an overlapping profile in EAC and GEJ, and across all clinical subtypes (**Figure 4, Extended Data Figure 6**), as further detailed in the **Supplementary Note**. The dataset included 179 cancer genes harboring driver alterations, including genes commonly implicated in EAC and gastric adenocarcinomas such as *TP53* (81.7%), *CDKN2A* (19.6%), and *KRAS* (13.8%)^9,11,38^.

**Figure 4.**
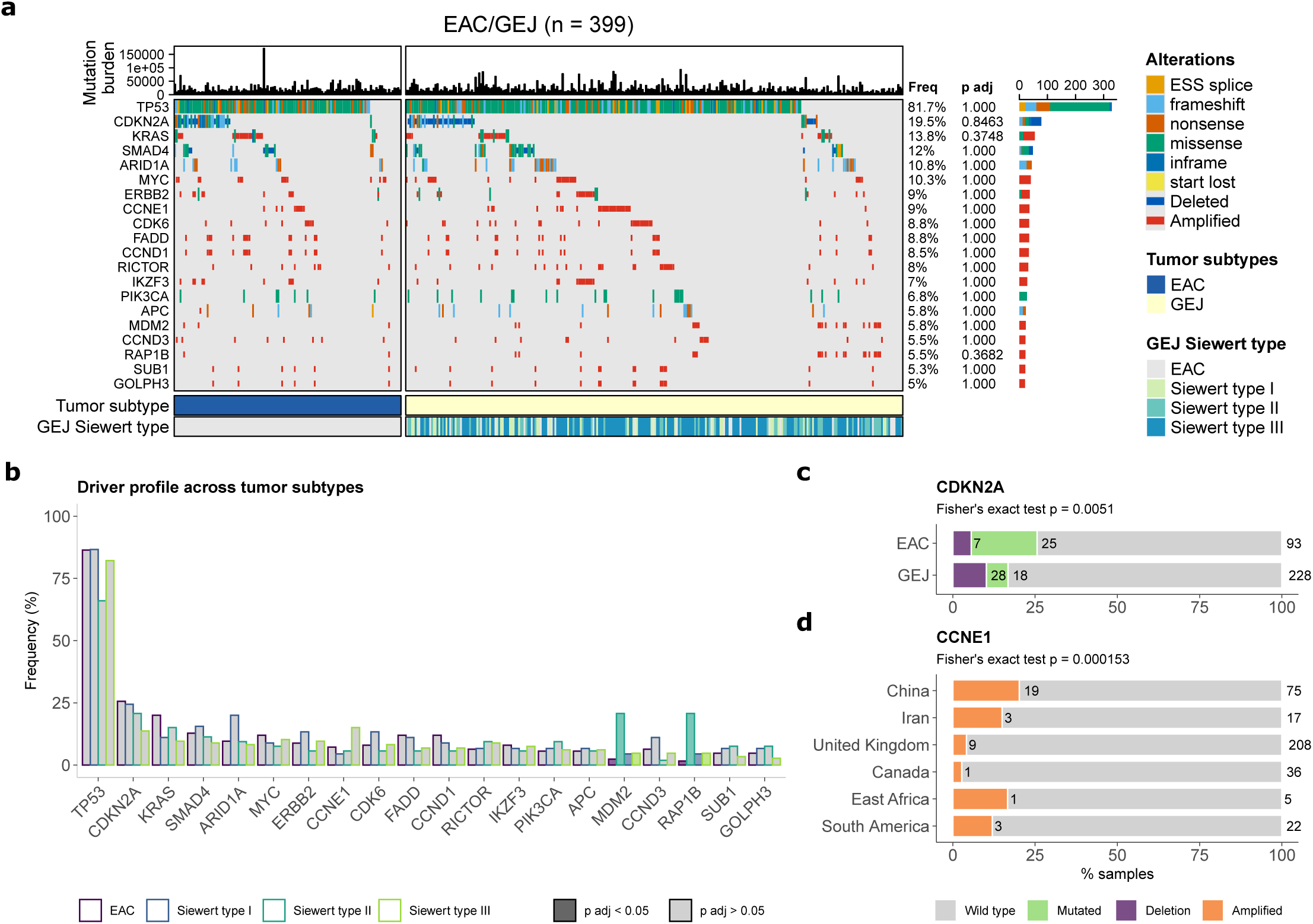
Homogeneous driver profile in EAC and GEJ. **a**, Driver mutations in 399 EAC/GEJ with available mutation and CN information, sorted by tumor subtype. Top, mutational burden per sample. Middle, presence of driver alterations per sample. Bottom, tumor subtype and Siewert type per sample. Frequency of mutations and q values from two-sided Fisher’s exact test comparing EAC and GEJ are displayed. **b**, Prevalence of driver mutations in driver genes across EAC and GEJ Siewert types. Genes with significantly different frequencies are colored. In **a-b**, genes with driver mutations or CN alterations in >5% of cases are shown. **c**, Frequency of CDKN2A mutations or deletions across EAC and GEJ samples. **d**, Frequencies of CCNE1 amplifications across geographical regions. The number of samples per category is indicated. Significance was assessed using Fisher’s exact test. In **a-d**, significance was corrected for multiple testing with the Bonferroni method.

### Reflux-related mutagenesis is associated with obesity in esophageal cases

We next investigated the mutational impact of lifestyle risk factors and EAC/GEJ-related exposures and identified an association between obesity at diagnosis and signatures SBS17a and SBS17b, proposed to be related to reflux-driven oxidative injury in EAC/GEJ^8^ (**Figure 5**). Both the signature burdens and frequencies of these signatures were significantly higher in obese patients compared to other BMI categories. However, stratification by tumor site revealed that this association was restricted to EAC cases, whereas no such association was observed in GEJ (**Figure 5a-c**). Multivariate logistic regression analysis confirmed that the enrichment of SBS17a/b signatures in obese EAC cases remains significant after correcting for potential confounders (**Figure 5d, Supplementary Table 10-Supplementary Table *12***). Consistent results were observed when examining historical obesity status prior to diagnosis, further reinforcing the link between obesity and bile acid reflux-related mutagenesis in EAC, which is not confounded by potential cancer-associated weight loss at diagnosis (**Supplementary Note**). These findings provide a molecular rationale for the link between obesity and EAC, suggesting that obesity may intensify the mutagenic insult to the lower esophagus, potentially by exacerbating gastroesophageal reflux, as proposed by epidemiological studies^1,39^.

**Figure 5.**
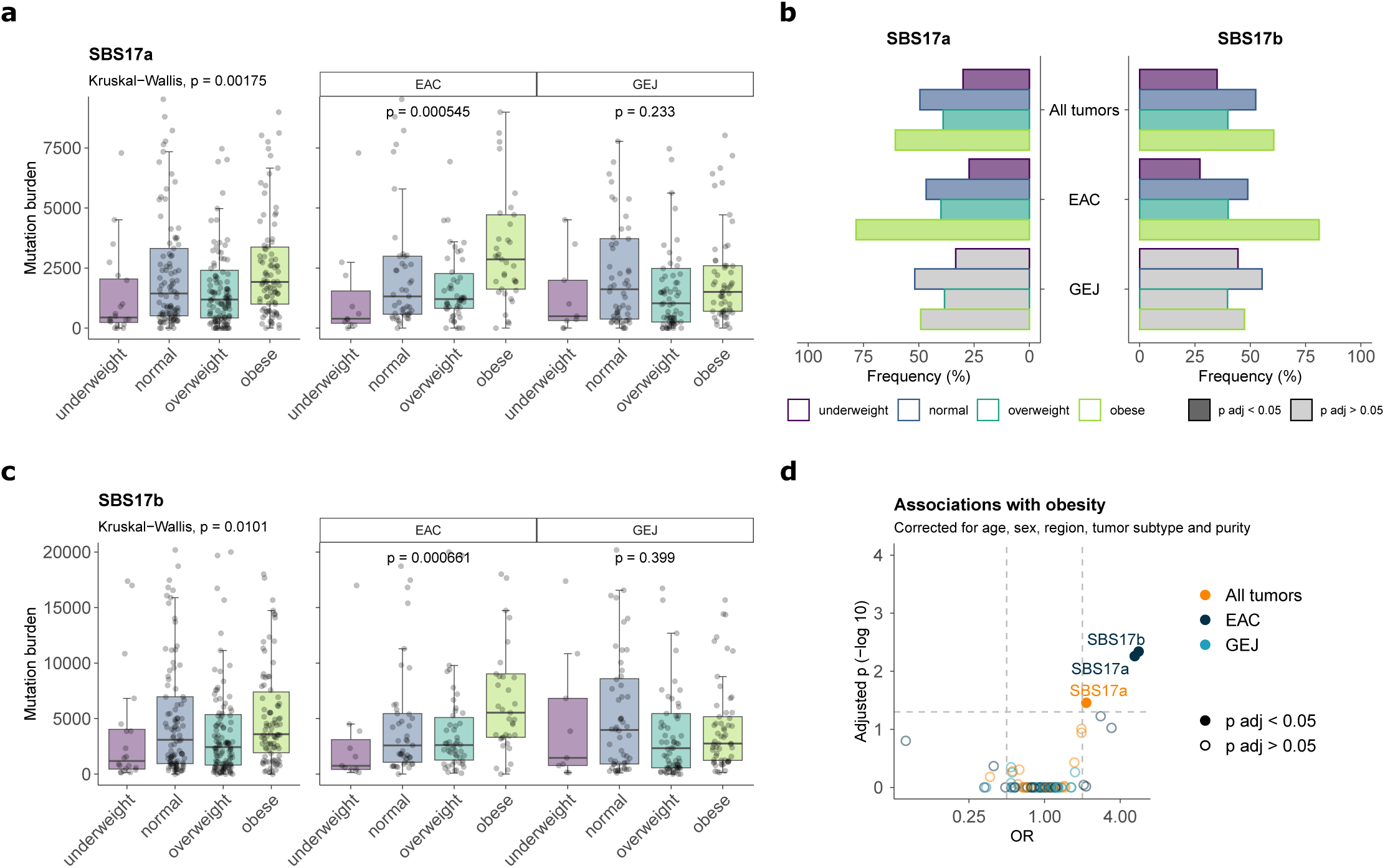
Associations of signatures with obesity. **a,c,** Mutational burdens for SBS17a (**a**) and SBS17b (**c**) by body mass index group, categorized as underweight (<18.5), normal (18.5 to 25), overweight (25 to 30), and obese (>30) (n = 486 biologically independent samples). Values from all tumors are depicted in the left panels, and across tumor subtypes in the right. The Kruskal–Wallis test (two-sided) was used to test for global differences. Box-and-whisker plots are in the style of Tukey. The line within the box is plotted at the median, while the upper and lower ends indicate the 25th and 75th percentiles. Whiskers show 1.5× IQR. The y axes were cut at 1.25× upper whisker for clarity. **b**, Frequencies of dichotomized SBS17a-b signatures across BMI categories for all tumors and by tumor subtype. Bars filled in gray indicate groups without significantly different frequencies across BMI categories. Significance was assessed using adjusted Fisher’s exact test. **d**, Associations between mutational signatures and obesity, measured by logistic regression. Signatures with significant associations (adjusted p-value < 0.05) in all tumors, EAC, or GEJ cases are colored. In **b,d**, P values were adjusted for multiple comparisons using the Bonferroni method based on the total number of signatures considered per variant type.

Self-reported clinical history of GERD was not associated with SBS17a/b signature positivity (**Supplementary Table 10-Supplementary Table *12***). However, the absence of symptom-based GERD does not exclude chronic reflux-driven injury, given that a substantial proportion of patients with BE and EAC have asymptomatic reflux, and GERD symptoms are frequently underreported^40,41^.

### No mutagenic effect related to tobacco and other EAC/GEJ-related exposures

None of the additional EAC/GEJ-related exposures evaluated, including tobacco smoking and alcohol consumption, exhibited a significant positive association with the presence of mutational signatures (**Supplementary Table 10-Supplementary Table *12***). Furthermore, no association between any of the risk factors and the spectrum of driver mutations or copy number alterations was identified (**Supplementary Table 2**,**Supplementary Table *16***). These data, coupled with the absence of signatures of exogenous factors, suggest that such exposures may contribute to EAC/GEJ through non-mutagenic mechanisms.

### Geographic variations in intracellular mutational processes

To understand whether the mutational processes driving EAC/GEJ are shared across geographical regions, we assessed the signature distribution across regions with different incidence rates^4,6^ (**Figure 6a,c**, **Supplementary Table 15**). We identified significant positive associations between age-standardized rate (ASR) of incidence and signatures related to intrinsic cellular processes, including SBS5, SBS8, SBS93, SBS103, and SBS111, alongside their associated ID signatures ID1, ID2, and ID14 (**Figure 6a-c**). Analysis of mutational signature distributions across regions revealed that SBS5, SBS93, and ID2 were significantly enriched in the high-incidence Shanxi region of China (**Figure 6d**). These regional differences were further mirrored in ancestry analyses based on genetic similarity, which confirmed a significant association between these signatures and individuals of East Asian descent (**Extended Data Figure 1d**). Conversely, we identified a significant inverse correlation between incidence and the ROS signature SBS18, as well as SBS102 (**Figure 6a-b**). The burdens of signatures SBS18 and SBS102 were particularly high in South America (**Figure 6b**, **Supplementary Table 15**) and among individuals of European ancestry (**Extended Data Figure 1d**, **Supplementary Table 13**).

**Figure 6.**
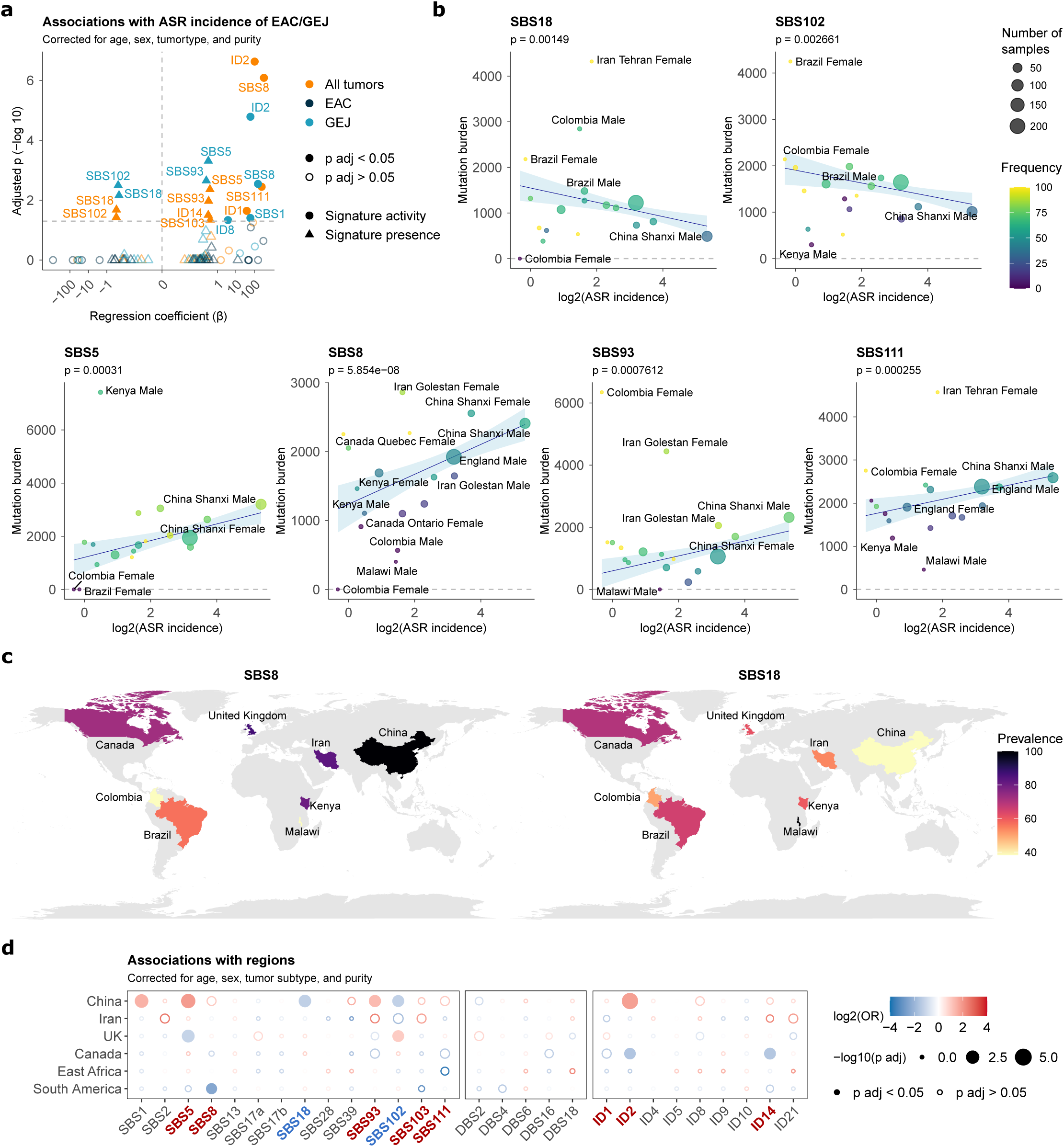
Geographic variation of signature profiles. **a-b**, Association of signature activities with ASR incidence of EAC and GEJ. Associations were evaluated using multivariate regression models with log2-transformed ASR incidence as an independent variable, adjusted for age at diagnosis, sex, tumor subtype, and tumor purity. For signatures detected in at least 75% of cases, linear regression was performed using signature activity as the dependent variable. Otherwise, logistic regression was performed using the binary presence/absence signature status as the dependent variable. **a**, Associations of signature activities with ASR incidence in the whole cohort, EAC, and GEJ. Signatures with significant associations (adjusted p-value < 0.05) are colored. Regression coefficients are displayed on a pseudo-log10 scale. **b**, Representative associations with SBS signatures in the whole EAC/GEJ cohort. Data are represented as average mutations attributed to SBS signatures per group. The number of cases per group and frequency of positive cases are indicated by size and color, respectively, and 95% confidence intervals are shown in clear blue. The P values shown correspond to the sample-level multivariate linear regression models used in **a**. **c**, Geographic distribution of the SBS8 and SBS18 mutational signatures. Countries were colored based on signature prevalence. **d**, Associations between mutational signatures and geographical region in EAC/GEJ, measured by logistic regression analysis. Regressions are adjusted for age of diagnosis, sex, tumor subtype, and tumor purity. Dots filled in color indicate significant associations (adjusted p-value < 0.05). Signatures that also showed significant positive and negative associations with ASR incidence are highlighted and colored on the x-axis (red and blue, respectively). In **a,d**, the Bonferroni method was used to adjust P values for multiple hypothesis testing, based on the total number of signatures considered per variant type.

Subgroup analysis by tumor subtype revealed that all geographic disparities were predominantly driven by GEJ, including positive associations between GEJ incidence and SBS5, SBS8, SBS93, and ID2, and negative associations with SBS18 and SBS102 (**Figure 6b**, **Supplementary Table 10-Supplementary Table *12***,**Supplementary Table *15***). In contrast, the mutational profile of EAC remained stable across all geographical regions and incidence rates. Overall, these results underscore global disparities in the EAC/GEJ mutation profile, particularly in GEJ.

Given the proposed role of *Helicobacter pylori* infection in the etiology of GEJ, described specifically in East Asia^17,42^, we evaluated the presence of *H. pylori* reads in tumor sequencing data (Methods, **Supplementary Table 14**). While *H.* pylori prevalence varied geographically, this could not explain the distinct mutational patterns observed in China (**Supplementary Note**, **Supplementary Table 10**Supplementary **Table *12***).

## DISCUSSION

The global distribution of EAC and GEJ is varied, with EAC incidence rising sharply over the last half-century^4,5^. By performing WGS of 520 tumors from geographical regions spanning four continents, our study revealed notable geographical differences in the substitution profiles of EAC/GEJ and provides insight into the mutational impact, or lack thereof, of established risk factors. It further implies shared mutagenic mechanisms underlying EAC and GEJ, challenging conventional clinical algorithms.

The anatomical boundary between gastric and esophageal adenocarcinomas remains debated. Previous genomic efforts have largely treated EAC and GEJ as separate entities or grouped Siewert type I-II GEJ with EAC and type III with gastric cancers^7,9–11^. Consequently, a direct, large-scale molecular comparison of these tumor subtypes has been lacking. Consistent with prior reports, both EAC and GEJ are characterized by high mutational burdens dominated by signatures SBS17a and SBS17b^7,10,11^. However, our analysis provides further granularity to their mutational landscape by identifying previously undetected signatures in EAC/GEJ, including SBS102, SBS103, and SBS111, originally described in colorectal and pan-cancer cohorts^13,34,43^. Our work reveals a fundamental similarity in mutational, driver, and copy-number profiles of EAC and tumors arising across the gastroesophageal junction, including those historically considered gastric in origin, suggesting that EAC and GEJ present a shared molecular landscape. Notably, only DNA repair signatures SBS111, ID1, and ID2 showed significantly different distributions across tumor subtypes, with higher activity in EAC and Siewert type I tumors than in type II and III GEJ. Our results reinforce earlier evidence of a shared molecular profile between EAC and chromosomally unstable gastric tumors^9^, and also align with previous observations describing a shared evolutionary molecular trajectory for all EAC tumors, regardless of a clinical history of BE^31^.

Signatures SBS17a and SBS17b were ubiquitously active across all EAC/GEJ subsites. These signatures have been identified as early mutational events^44,45^, and have been documented in BE^11^, overall positioning them as a key event for malignant transformation in EAC/GEJ. This contrasts with gastric cancers, which exhibit lower absolute SBS17 activity according to previous studies^38,46^, suggesting that the mutational profile in GEJ differs from the distal stomach. Reflux has been proposed as a likely cause of SBS17a/b in EAC/GEJ, and experimental studies have shown that exposure to bile and gastric acids increases the oxidative damage to guanine^47^, which could generate the characteristic T>G transversions that define the SBS17^23^. We observed an enrichment of SBS17a/b signatures in tumors from obese individuals. This aligns with the clinical observation that high body mass index may exacerbate reflux-mediated tissue damage, likely by increasing intra-abdominal pressure and subsequent chronic acid exposure^1,39^. However, other indirect obesity-related mechanisms independent of reflux could also contribute to this mutation profile^48^. While obesity is an established driver of EAC, its link to GEJ risk is less certain^17–20^, aligning with our results showing an association of SBS17a/b signatures with obesity only in EAC cases. A potential limitation of this analysis is the lack of abdominal obesity measures, which are more strongly associated with EAC risk than BMI alone^39^. Furthermore, SBS17a/b signatures also occur in other tumor types, including colorectal and breast cancer, albeit as late clonal events^13,37^. Thus, other oxidizing mechanisms besides reflux must trigger SBS17a/b signatures. For instance, fluorouracil chemotherapy has been reported to elicit SBS17a/b signatures in treated patients^24^.

Despite tobacco being a well-known mutagen, we found no mutational footprint attributable to tobacco smoking in EAC/GEJ tumor genomes, as well as no distinct driver alterations. Specifically, tobacco-related signatures observed in other cancer types (e.g., SBS4, SBS92, SBS100, and SBS109)^7,15,49,50^ were absent from the EAC/GEJ mutational spectra, and smoking showed no association with other mutational signatures. Consistent with findings in esophageal squamous cell carcinoma, where tobacco signatures were found only at low burdens^14^, these results suggest that tobacco and other exposures lead to EAC/GEJ development through non-mutagenic mechanisms. This observation further highlights the tissue-specific impact of tobacco throughout the aerodigestive tract^15^. One plausible explanation is that tobacco acts primarily as a tumor promoter in these tissues, consistent with recent studies supporting a promotional role for tobacco in other cancer types^51,52^. Similarly, while alcohol is a known mutagen in several cancers, alcohol-related signatures (e.g., SBS16) were not detected, reinforcing epidemiological evidence that this exposure may not be a risk factor for EAC/GEJ^21,22^.

Finally, our analysis reveals significant mutational disparities across geographical regions, particularly in GEJ from the high-incidence Shanxi region in China. As observed in other tumor types studied within the Mutographs framework, specifically esophageal squamous cell carcinoma and pancreatic cancer^14,53^, geographic differences in incidence are not explained by variation in exogenous mutational signatures, which further supports the hypothesis that key environmental exposures lack a direct mutagenic effect. However, the EAC/GEJ mutational landscape is uniquely distinguished by geographic variation in signatures related to internal cellular processes and DNA repair infidelity. These intrinsic signatures correlate with EAC/GEJ incidence rates, yet are unrelated to the risk factors examined. These observations suggest that elevated cancer risk in certain populations may arise from dysregulation of multiple intrinsic cellular processes rather than the direct effect of an exogenous mutagen. Such global variations may reflect ancestry-specific susceptibilities, or alternatively, collateral mutagenesis from multiple sources of exogenous DNA damage, a recently proposed mechanism for SBS5^54^. Interestingly, two signatures exhibit geographic patterns similar to those observed in colorectal cancer. SBS103 is positively associated with the incidence of both EAC/GEJ and colorectal cancer^13^, while the enrichment of SBS102 in European and South American EAC/GEJ mirrors its reported prevalence in colorectal cancers from Colombia^13^. Whether these signatures reflect endogenous or environmental exposures remains to be determined.

Our findings also provide insight into the evolutionary history of EAC/GEJ. Signatures associated with EAC/GEJ incidence, including SBS8 and SBS111, are present among late clonal mutations in EAC/GEJ, while they are absent in healthy esophagus and gastric tissues^11,55,56^, suggesting that they likely manifest during neoplastic progression rather than as an initiating event. In contrast, SBS17a/b activity remains uniform across regions, contrary to previous reports in a small EAC cohort from China^57^. Together, these observations suggest that the core early-stage mutagenic process driving EAC/GEJ is uniform across regions, while geographic differences emerge at later stages of tumor evolution through variations in intrinsic cellular and DNA repair processes.

In summary, this work provides a unified genomic landscape for EAC and GEJ and elucidates the molecular forces driving these malignancies, including an association between reflux-related signatures and obesity in EAC, as well as differences in the geographic distribution of patterns of intrinsic mutagenesis. Ultimately, our findings suggest that EAC and GEJ present a shared molecular landscape, supporting the inclusion of molecular characteristics, and not only anatomical boundaries, in the decision-making for staging systems and prevention strategies.

## METHODS

### Recruitment of cases and informed consent

The International Agency for Research on Cancer (IARC)/World Health Organization (WHO) coordinated participant recruitment, comprising 12 collaborating centers from the eight participating countries in Africa, America, Asia, and Europe (**Supplementary Table 17**). Countries with n<5 samples were grouped into regions for downstream analyses (South America, including Brazil and Colombia, and East Africa, including Kenya and Malawi). Inclusion criteria for patients were ≥18 years of age (ranging from 32 to 87 years, with a mean of 66 and standard deviation of 10 years), confirmed diagnosis of primary EAC/GEJ, and no prior treatment. Written informed consent was obtained for all participants. Patients were excluded if they had any condition that could interfere with their ability to provide informed consent or if there were no means of obtaining adequate tissues as per protocol requirements. Ethical approvals were first obtained from each local research ethics committee and federal ethics committee when applicable, as well as from the IARC Ethics Committee.

### Bio-specimens and data collection

Dedicated standard operating procedures, following guidelines from the International Cancer Genome Consortium, were designed by the IARC/WHO to select adequate case series with complete biological samples and exposure information as described previously^12,14^ (**Supplementary Table 17**). In brief, anthropometric measures were taken for all case series included, along with relevant information regarding medical and family history. All biological samples from retrospective cohorts were collected using rigorous, standardized protocols and fulfilled the required standards of sample collection defined by the IARC/WHO for sequencing and analysis. Retrospective case series were included after examination of their respective recruitment protocols to ensure the availability of necessary biological samples based on standard operating procedures, following guidelines from the International Cancer Genome Consortium, and also based on the collection of relevant exposure history based on a comparison of validated epidemiological questionnaires from each specific region. All patient-related data, as well as clinical, demographical, lifestyle, pathological, and outcome data, were pseudonymized locally using a dedicated alphanumeric identifier system before being transferred to the IARC/WHO central database.

### Expert pathology review

Original diagnostic pathology departments provided diagnostic histological details of contributing cases through standard abstract forms, together with a representative hematoxylin–eosin-stained slide of formalin-fixed paraffin-embedded tumor tissues whenever possible. The IARC/WHO centralized the entire pathology workflow and coordinated a centralized digital pathology examination of frozen tumor tissues collected for the study, as well as formalin-fixed paraffin-embedded sections when available, via a web-based report approach and a dedicated expert panel, following standardized procedures^58^. A minimum of 50% viable tumor cells was required for eligibility for whole-genome sequencing.

### DNA extraction

A total of 904 patients with primary esophageal and esophagogastric junction adenocarcinomas were enrolled in the study. Of these, 349 samples (39%) were excluded due to insufficient viable tumor cells (pathology level). Extraction of DNA from fresh frozen tumors and matched blood or normal samples was centrally conducted at IARC/WHO, except for cases from the United Kingdom (n = 306), for which DNA extraction was conducted at Sanger. Germline DNA was extracted from either blood samples (554 cases) or normal tissues (1 case) using previously described protocols and methods^14^. A total of 555 cases were sent to the Sanger Institute for DNA sequencing.

### Whole-genome sequencing

Fluidigm SNP genotyping with a custom panel was performed to ensure that each pair of tumor and matched normal samples originated from the same individual. Whole-genome sequencing (150 bp paired-end) was performed on the NovaSeq 6000 platform with a target coverage of 40x for tumors and 20x for matched normal tissues. All sequencing reads were aligned to the GRCh38 human reference genome using Burrows-Wheeler-MEM (v.0.7.16a and v.0.7.17)^59^. A standard set of post-sequencing quality criteria was applied for metrics, including total coverage, evenness of coverage, and contamination. Cases were excluded if coverage was below 30x for tumors or 15x for normal tissue. For evenness of coverage, the median over mean coverage (MoM) score was calculated. Tumors with MoM scores outside the range of values determined by previous studies to be appropriate for whole-genome sequencing (0.92–1.09) were excluded^60^. Conpair^61^ was used to detect contamination, and any tumor or normal sample with a value above 3% was excluded^61^. A total of 520 cases passed all criteria and were included in subsequent analysis.

### Somatic variant calling

A standard analysis pipeline (https://github.com/cancerit) was used to perform variant calling for copy number variants (CNVs; ASCAT^62^ and Battenberg^63^, when tumor purity allowed), SNVs (cgpCaVEMan^64^), indels (cgpPindel^65^) and structural rearrangements (BRASS, https://github.com/cancerit/BRASS). CaVEMan and BRASS were run using the copy number profile and purity values determined from ASCAT when possible (complete pipeline, n = 428), or using copy number defaults and an estimate of purity obtained from ASCAT-Battenberg when tumor purity was insufficient to determine an accurate copy number profile (partial pipeline, n = 92). For SNVs, additional filters (ASRD ≥ 140 and CLPM = 0) were applied in addition to the standard PASS filter to remove potential false positive calls. To further exclude the possibility of caller-specific artifacts being included in the analysis, a second variant caller, Strelka2, was run for SNVs and indels^14,66^, with variants called by both the Sanger variant-calling pipeline and Strelka2 included in subsequent analyses. All downstream analyses were performed on a subset of 486 samples with no evidence of DNA repair deficiencies or prior chemotherapy treatment (**Supplementary Note**).

### Generation of mutational matrices

Mutational matrices for SBS, DBS, and indels were generated using SigProfilerMatrixGenerator with default options (v1.2.30)^67^.

### Mutational signature extraction

Multiple methods were used to extract mutational signatures. The primary extractions were performed using SigProfilerExtractor^35^, based on non-negative matrix factorization. mSigHdp^68^, based on hierarchical Dirichlet process mixture models, was used as a second method to validate the de novo mutational signatures extracted. SigProfilerExtractor (1.1.24) was run using nndsvd_min initialization (NMF_init=‘nndsvd_min’) for 1–20 signature solutions and 500 non-negative matrix factorization (NMF) replicates. For SBS, mutational signatures were extracted in both SBS1536 and SBS288 contexts.

Both results were similar (**Supplementary Note**), with the SBS1536 results taken forward for the final analysis (**Supplementary Table 3**). Signatures were extracted using SigProfilerExtractor in the following contexts for other variant types: DBS78 for DBS, and ID83 (**Supplementary Table 4**-**Supplementary Table *5***). mSigHdp extractions were performed using the suggested parameters and using the country of origin to construct the hierarchy for SBS96 and ID83 contexts. A comparison of the SigProfilerExtractor and mSigHdp results can be found in the **Supplementary Note**.

### Decomposition of mutational signatures

After de novo extraction was completed, SigProfilerAssignment^69^ v0.1.4 was used to decompose the de novo extracted SBS, ID, and DBS mutational signatures into COSMIC v3.6^70^ (**Supplementary Table 6**). When possible, SigProfilerAssignment matched each de novo extracted mutational signature to a set of previously identified COSMIC v3.6 signatures. This step is important as it allows the detection of de novo signatures that are made up of multiple reference signatures that have not separated during the extraction process. Decompositions were performed using both default and optimized settings. To optimise the decomposition, the exclude_signature_subgroups parameter was used to exclude artefact signatures, ultraviolet signatures, lymphoid signatures, mismatch repair deficiency signatures, polymerase deficiency signatures, homologous recombination deficiency signatures, base excision repair deficiency signatures, and treatment signatures. The optimised decomposition results were used for all signatures except one (**Supplementary Note**).

### Attribution of mutational signatures to individual samples

The activities of the de novo and COSMIC signature panels were attributed to each sample using MSA^71^ (v2.0) (**Supplementary Table 7Supplementary Table *8***). For decomposed attributions, only the panel of COSMIC reference signatures identified during the decomposition of mutational signatures was included. A conservative approach was used for MSA attributions using the (params.no_CI_for_penalties=False) option for the calculation of optimum penalties. Pruned attributions were used for the final analysis, where confidence intervals were applied to each attributed mutational signature, and any signature activity with a lower confidence limit equal to 0 was removed.

### Driver mutations

Driver mutations in EAC/GEJ were identified using the following methods. First, the normalized ratio of non-synonymous to synonymous mutations (dN/dS) was used to identify genes under positive selection^56^. Results were calculated both for the whole genome (q < 0.01) and with restricted hypothesis testing for a panel of 369 known cancer genes. Variants in any gene identified as under positive selection in global dN/dS or in the 369-cancer gene panel were considered as potential driver mutations and were then classified as likely drivers if they met any of the following criteria: (1) truncating mutations in genes annotated as tumor suppressors, (2) mutations annotated as likely or known oncogenic in MutationMapper, (3) truncating variants in genes with selection (q < 0.05) for truncating mutations assumed to be tumor suppressors and thus likely drivers, (4) missense variants in all genes under positive selection and with dN/dS ratios for missense mutations above five (assuming four of every five missense mutations are drivers) labeled as likely drivers or (5) in-frame indels in genes under significant positive selection for in-frame indels. The Cancer Gene Census (https://cancer.sanger.ac.uk/census) and the Cancer Genome Interpreter tool (https://www.cancergenomeinterpreter.org) were used to annotate potential drivers with the mode of action. Missense mutations were assessed using the MutationMapper tool (http://www.cbioportal.org/mutation_mapper).

### Copy number profile

The copy number profiles were investigated in a subset of cases with available copy number data (n = 399). Driver copy number alterations were defined as cancer-related alterations in the COSMIC cancer gene census (v102) reported as amplified or deleted, or alterations in COSMIC genes reported as CN drivers in EAC and GEJ in previous studies^10,72^. Homozygous deletions were defined as copy number = (0, 0), and amplifications as copy number > 2 × ploidy + 1. Unsupervised clustering analysis of the copy number counts was performed using Euclidean distance and Ward’s agglomerative procedure.

### Evolutionary analysis

DPClust^63^ (v2.2.8) was used to infer the clonal structure of 399 samples with available Battenberg copy-number profiles (complete pipeline) and mutational signature information. DPClust3p (v1.0.8) was applied for data pre-processing. MutationTimeR (v1.00.1)^37^ was run on the DPClust output to assign SBS and ID mutations to either early clonal, late clonal, subclonal or unspecified clonality status (not assigned). Samples with at least 256 early clonal or late clonal SBS, and at least 100 early clonal or late clonal ID were retained for downstream analysis (n = 362 for SBS and 316 for ID analyses). Annotated VCFs were subsequently split into early clonal and late clonal subsets and analyzed separately. MSA^71^ (v2.0) was run on each subset to identify the mutational processes active in the early clonal and late clonal mutations for each sample, using pruned signature attributions. Differences between the early and late relative activity of each mutational signature were assessed using a Wilcoxon signed-rank test, and P values were corrected across signatures using the Benjamini– Hochberg procedure (q value).

### *Helicobacter pylori* detection

To identify microbial reads mapping to *Helicobacter pylori*, low-quality reads were first filtered using fastp (v0.24.0)^73^. Remaining human reads were removed by excluding sequences that mapped to the human reference genomes GRCh38, T2T-CHM13 v2.0^74^, and the Human Pangenome Reference Consortium (HPRC) 47-genome pangenome assembly^75^. The resulting high-quality, non-human reads were analyzed using two orthogonal taxonomic classification approaches to detect *H. pylori*. First, taxonomic abundance estimation was performed using Bracken (v2.9)^76^ with the KrakenUniq MicrobialDB database (release 8 August 2023; https://benlangmead.github.io/aws-indexes/k2). Second, taxonomic profiling was conducted using MetaPhlAn (v4.0.6)^77^ with the mpa_vJun23_CHOCOPhlAnSGB_202307marker gene database (June 2023 release). Using predefined detection thresholds (Bracken ≥10 reads; MetaPhlAn ≥0.1 relative abundance), both approaches demonstrated high concordance in *H. pylori* detection across tumor and normal samples. Overall agreement was 88.3% (κ = 0.31).

### Genomic ancestry and admixture analyses

Germline SNVs and IDs were derived from whole-genome sequencing from the normal paired material for each individual using Strelka2 with standard quality control criteria^66^. Variants were restricted to HapMap Phase 3 SNPs and filtered to retain only biallelic SNPs with minor allele frequency ≥1%, genotype missingness ≤5%, and Hardy–Weinberg equilibrium p ≥1×10⁻⁸. After pruning for long-range linkage disequilibrium regions and linkage disequilibrium (r2 < 20% within a 50 kb window), 135,987 independent variants remained in genotype data. Reference genotype data from the 1000 Genomes Project^78^ from five superpopulations (European, African, East Asian, South Asian, and Amerindian, n = 3,202 individuals) were filtered using the same criteria and merged with the EAC/GEJ dataset based on the pruned set of variants present in both datasets. ADMIXTURE analysis was performed on the combined dataset, with k = 5 ancestral populations selected based on model fit, and ancestry proportions were estimated for each sample (**Supplementary Table 18**). To complement these results, principal component analysis was run on the same merged dataset (**Supplementary Note**). Samples were assigned to a superpopulation if their corresponding ancestry proportion exceeded 0.7. This threshold was determined empirically by maximizing classification accuracy in the 1000 Genomes reference data.

### Regressions and associations with signatures

Signature attributions were dichotomized into presence and absence based on confidence intervals, with presence defined as both lower and upper limits being positive and absence as the lower limit being zero. Signatures present in at least 75% of cases (signatures SBS1, SBS8, categorizing SBS17a, SBS17b, SBS111, ID1, ID2, and ID8) were dichotomized into above and below the median of attributed mutation counts. The binary attributions served as dependent variables in logistic regression models, with relevant risk factors or epidemiological characteristics treated as factorized independent variables. Regressions with variables presenting data separation were performed using Firth’s penalized logistic regression. To adjust for confounding factors, sex, age of diagnosis, tumor subtype (EAC vs GEJ), region, and tumor purity were added as covariates in the regressions. The region variable was dichotomized as “China vs others” for whole-cohort and GEJ-specific analyses, and as “UK vs others” in EAC-specific analyses. P-values were corrected for multiple testing using the Bonferroni method.

The relationship between signature activity and ASR of incidence was evaluated on a per-sample basis, aligning with previous work^15,79^. Regional-level ASRs were obtained from Cancer Incidence in Five Continents (CI5), where available^6^; otherwise, country-level ASRs from the Global Cancer Observatory (GLOBOCAN) were used (Brazil, Iran, and Malawi)^4^. Incidence rates for C15.0 adenocarcinomas of the esophagus were used for EAC, and rates for C16.0 gastric cancers were used for GEJ. For signatures present in at least 75% of cases, linear regression was performed using continuous signature attribution as the dependent variable. For signatures present in <75% of cases, logistic regression was performed using the binary presence/absence signature status as the dependent variable as detailed above. The log2-transformed ASR served as the independent variable: combined EAC and GEJ ASRs were used for whole-cohort analyses, while tumor-type-specific ASRs were used for subgroup analyses in EAC and GEJ. Regressions were adjusted for age at diagnosis, sex, tumor subtype, and tumor purity.

## DATA AVAILABILITY

Whole genome sequencing data and patient metadata are being deposited in the European Genome-phenome Archive (EGA) associated with the study EGAS00001003702, including CRAM files for all EAC/GEJ cases included in the final analysis, consensus SNV and indel variant-calling files, CNV calling, and patient metadata. All other data is provided in the accompanying Supplementary Tables, including mutation spectra of the extracted signatures, as well as signature activities, driver mutations, and CNV in the EAC/GEJ dataset.

## CODE AVAILABILITY

All algorithms used for data analysis are publicly available with repositories noted within the respective method sections and in the accompanying reporting summary. Code used for regression analysis and figures is available at GitLab https://gitlab.com/Mutographs/mutographs-eac-gej.

## ACKNOWLEDGEMENTS

The authors thank the IARC General Services, including the Laboratory Services and Biobank team led by Z. Kozlakidis and the Section of Support to Research overseen by C. Mehta; L. O’Neill, K. Roberts, K. Smith, S. Austin-Guest and the staff of Sequencing Operations at the Wellcome Sanger Institute for their contribution to sequencing; the work of all other collaborators in the Mutographs project who participated in the recruitment of patients in all centres; and all the patients involved in this study and their families. The authors also thank Mimi McCord for her contributions as patient advocate during the Mutographs project. The authors thanks Nicola Grehan, Ginny Devonshire, Barbara Nutzinger and Adam Freeman for their contributions to the OCCAMS study. This work was delivered as part of the Mutographs team, supported by the Cancer Grand Challenges partnership funded by Cancer Research UK (C98/A24032). Work performed by the Section of Support to Research at the IARC/WHO was supported by regular budget funding. The views expressed in this article by authors identified as IARC/WHO personnel are their own and do not necessarily represent the decisions, policies, or views of the IARC/WHO. This work was supported by the Wellcome Trust grants 206194 and 220540/Z/20/A. The OCCAMS study was supported by Cancer Research UK [A15874, A22720, A22131]. This work uses data provided by patients and collected by the NHS as part of their care and support. The OCCAMS data disclosed in the publication may only be used by not-for-profit organizations and only for academic, teaching, clinical care and educational purposes. Work at CNIO was also supported by the Agencia Estatal de Investigación (AEI/10.13039/501100011033), Ministerio de Ciencia, Innovación y Universidades, and co-funded by the European Regional Development Fund (ERDF-EU; PID2024-158474NA-I00). P.G.G. and M.D.-G. were awarded fellowships within the “Generación D” initiative, Red.es, Ministerio para la Transformación Digital y de la Función Pública, for talent attraction (C005/24-ED CV1), funded by the European Union NextGenerationEU funds, through PRTR. This research was supported by the Intramural Research Program of the National Institutes of Health (NIH). The contributions of the NIH author (A.M.G.) are considered Works of the United States Government. The findings and conclusions presented in this paper are those of the author and do not necessarily reflect the views of the NIH or the U.S. Department of Health and Human Services. For the purpose of Open Access, the author has applied a CC BY public copyright license to any Author Accepted Manuscript version arising from this submission. The designations employed and the presentation of the material in this publication, in particular in Fig. 1, do not imply the expression of any opinion whatsoever on the part of the authors or their institutions concerning the legal status of any country, territory, city, or area or of its authorities, or concerning the delimitation of its frontiers or boundaries.

## CONTRIBUTIONS

L.B.A., P.B., M.R.S, and S.P. conceived, designed, and supervised the study. L.T., S.M., J.P., H.Z., A.A., C.N., S.F., A.N., P.G.G., V.G., T.C., S.C., E.T., and M.D.G. conducted data analysis. B.A-A designed and oversaw the pathology workflow and performed pathology evaluation. P.C., C.L., and C.C. carried out sample manipulation. M.P.C., C.D., F.F.C., L.F., R.C.F., A.M.G., A.H., R.M., D.M., R.M.R., F.R.P., G.R., and J.Y. led or facilitated patient and sample recruitment. S.F., S.C., and E.T. performed data generation. A.C.d.C., and L.H. carried out scientific project management. L.T. and S.M. jointly contributed and were responsible for overall scientific coordination. L.T., S.M., L.B.A., P.B., M.R.S., and S.P. wrote the manuscript, with contributions from all other authors. All authors read and approved the final manuscript.

## EXTENDED DATA FIGURE LEGENDS

**Extended Data Figure 1.**
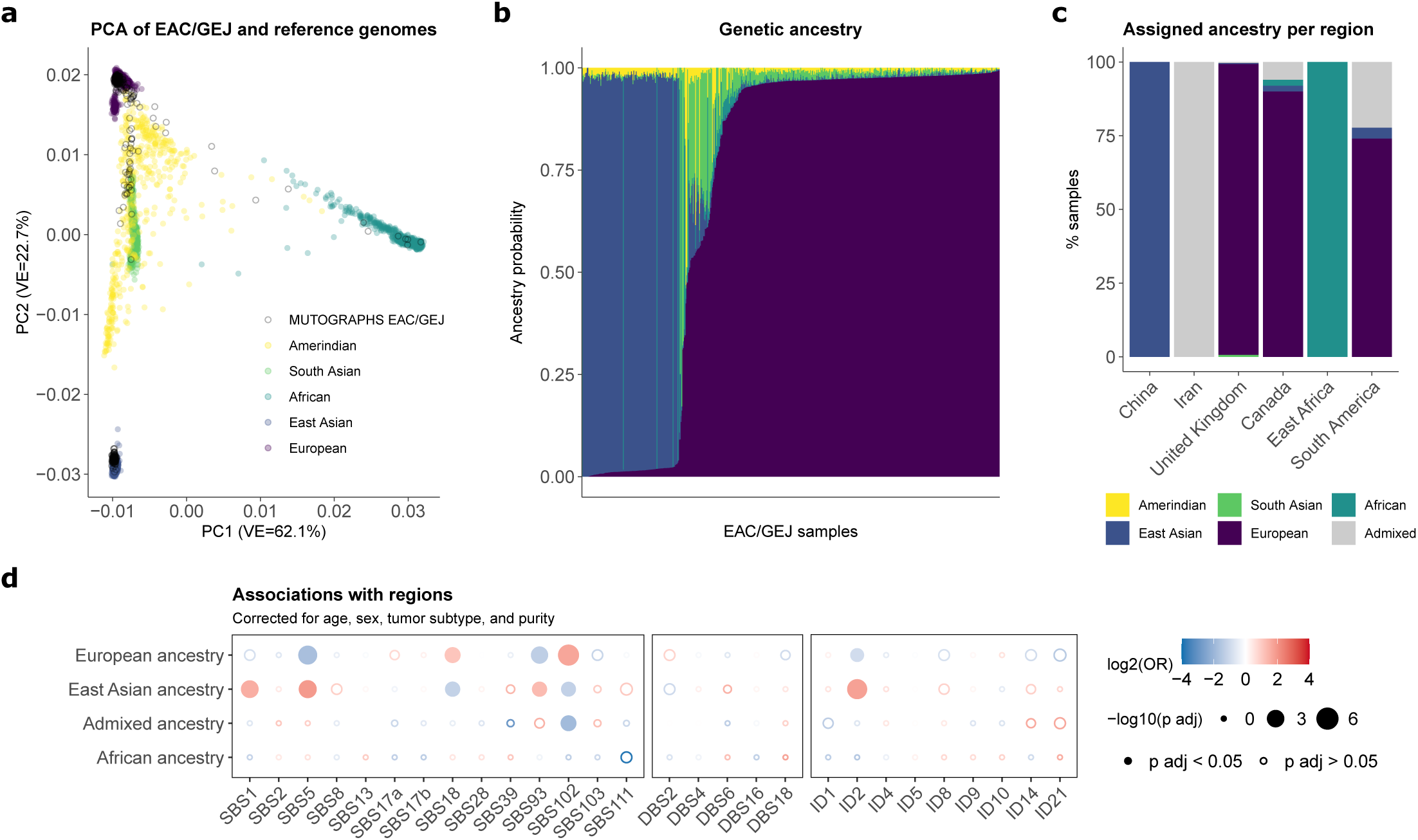
Signature distribution across ancestry groups. **a,** Scatter plots of principal components (PC) PC1, and PC2 based on genotype data showing the genetic ancestry of the EAC/GEJ cohort across geographical regions. **b**, Ancestry admixture in the EAC/GEJ Mutographs cohort. **c**, Relative proportions of samples assigned to each ancestry group. **d**, Associations between mutational signatures and ancestry groups in EAC/GEJ, measured by logistic regression analysis. The Amerindian ancestry group was discarded from the analysis due to insufficient sample size (n=2). Dots filled in color indicate significant associations (adjusted p-value < 0.05). Regressions were corrected for age of diagnosis, sex, tumor subtype, and tumor purity. The Bonferroni method was used to adjust P values for multiple hypothesis testing, based on the total number of signatures considered per variant type.

**Extended Data Figure 2.**
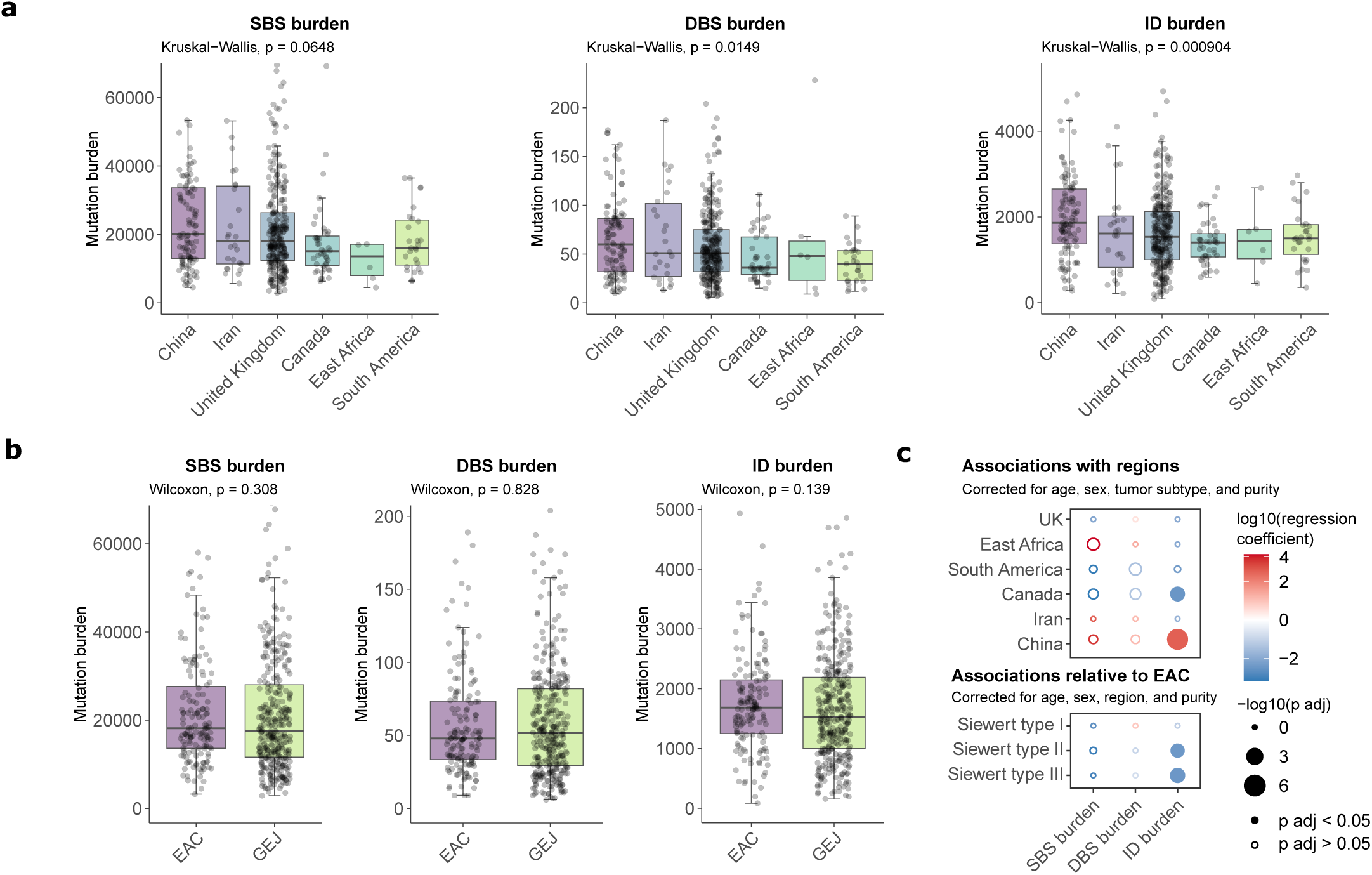
Mutational burdens in EAC/GEJ. **a-b**, Mutational burdens for SBSs, DBSs, and small ID burdens by geographical region (**a**) and tumor subtype (**b**). Kruskal–Wallis test (two-sided) was used to test for global differences. Box-and-whisker plots are in the style of Tukey. The line within the box is plotted at the median, while upper and lower ends indicate 25th and 75th percentiles. Whiskers show 1.5× interquartile range (IQR). **c**, Linear regression analysis of the association between mutation burdens and geographical regions or GEJ Siewert type relative to EAC (reference). Dots filled in color indicate significant associations (adjusted p-value < 0.05). Covariates included in the regressions are indicated. The Bonferroni method was used to adjust P values for multiple hypothesis testing.

**Extended Data Figure 3.**
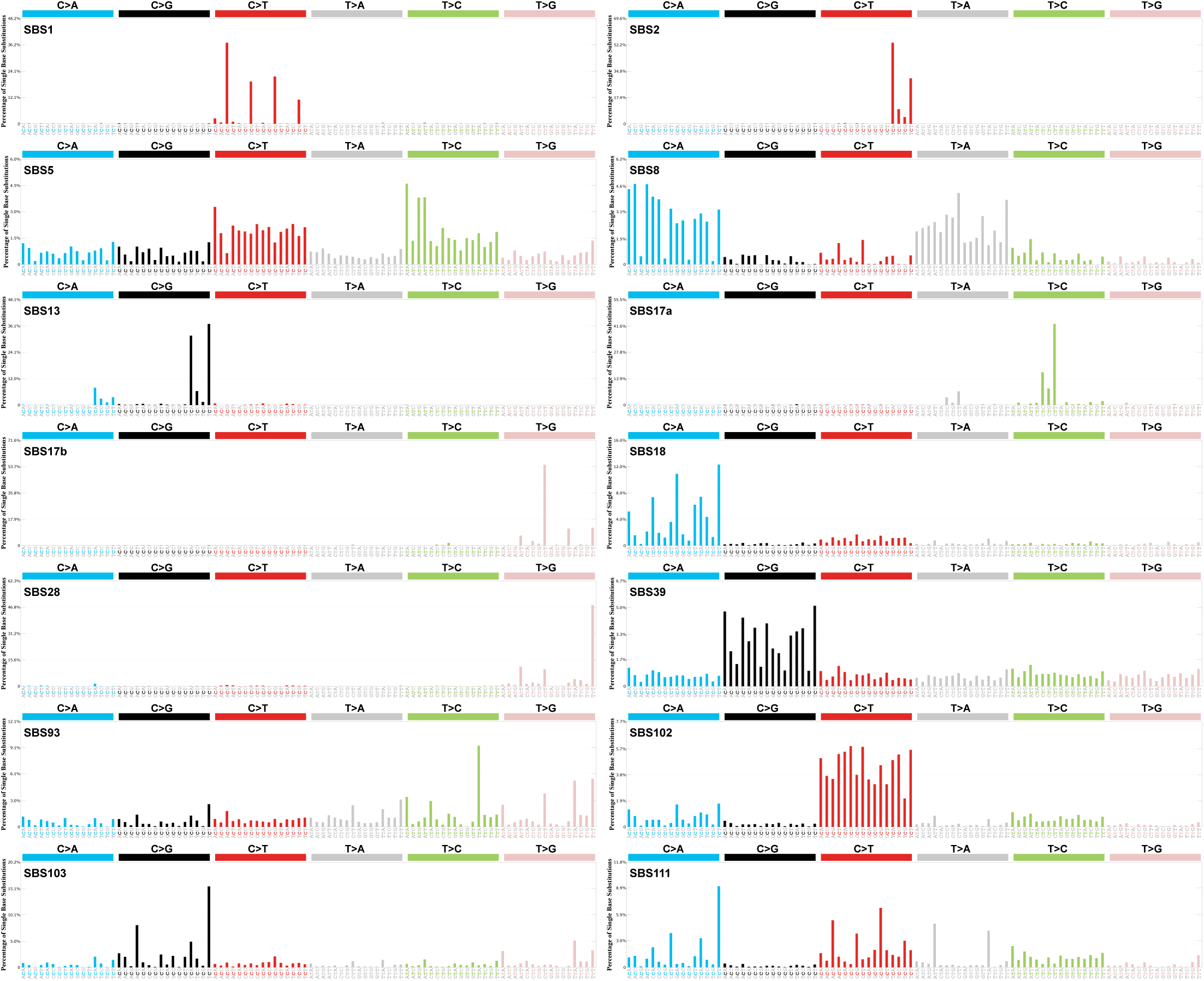
SBS signature decomposition. Decomposed SBS signatures, including reference COSMIC signatures.

**Extended Data Figure 4.**
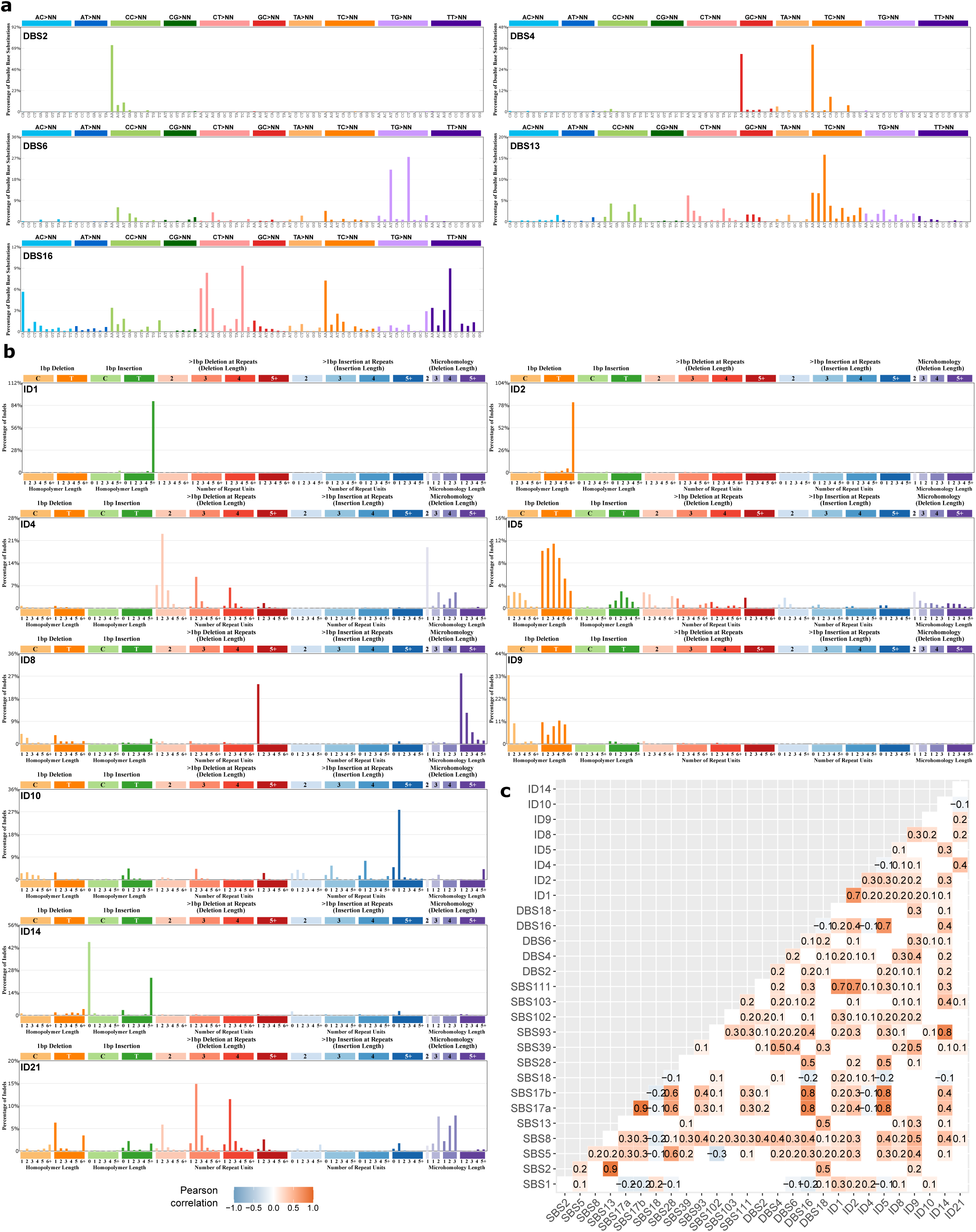
DBS and ID signature decomposition. **a,b,** Decomposed DBS (**a**) and ID (**b**) signatures. **c,** Correlations amongst mutational signatures. Pearson correlation coefficients for each significant comparison are indicated.

**Extended Data Figure 5.**
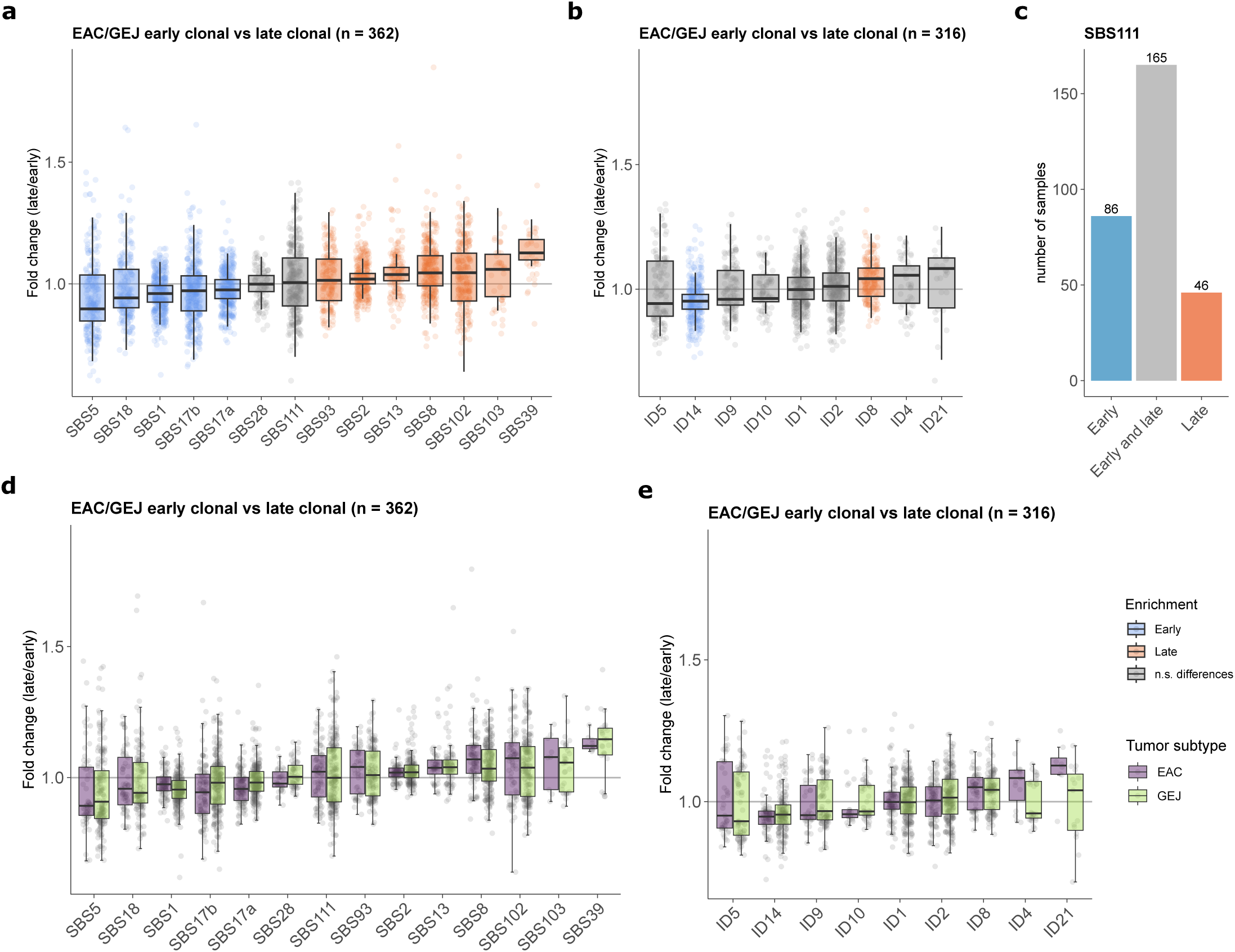
Evolutionary analysis. Fold change of the relative contribution per sample of each signature between early clonal and late clonal mutations. Signatures were sorted by median fold change. **a-b,** SBS (**a**) and ID (**b**) signatures with significantly greater relative burdens in early or late clonal stages are indicated in color (q < 0.05). Significance was assessed using a two-sided Wilcoxon signed-rank test, and p-values were corrected using the Benjamini–Hochberg procedure (q value). c, Number of EAC/GEJ samples presenting signature SBS111 in early clonal mutations, late clonal mutations, and both. **d-e**, Fold change contribution for SBS (**d**) and ID (**e**) signatures in early clonal and late clonal mutations in cases sorted by tumor subtype.

**Extended Data Figure 6.**
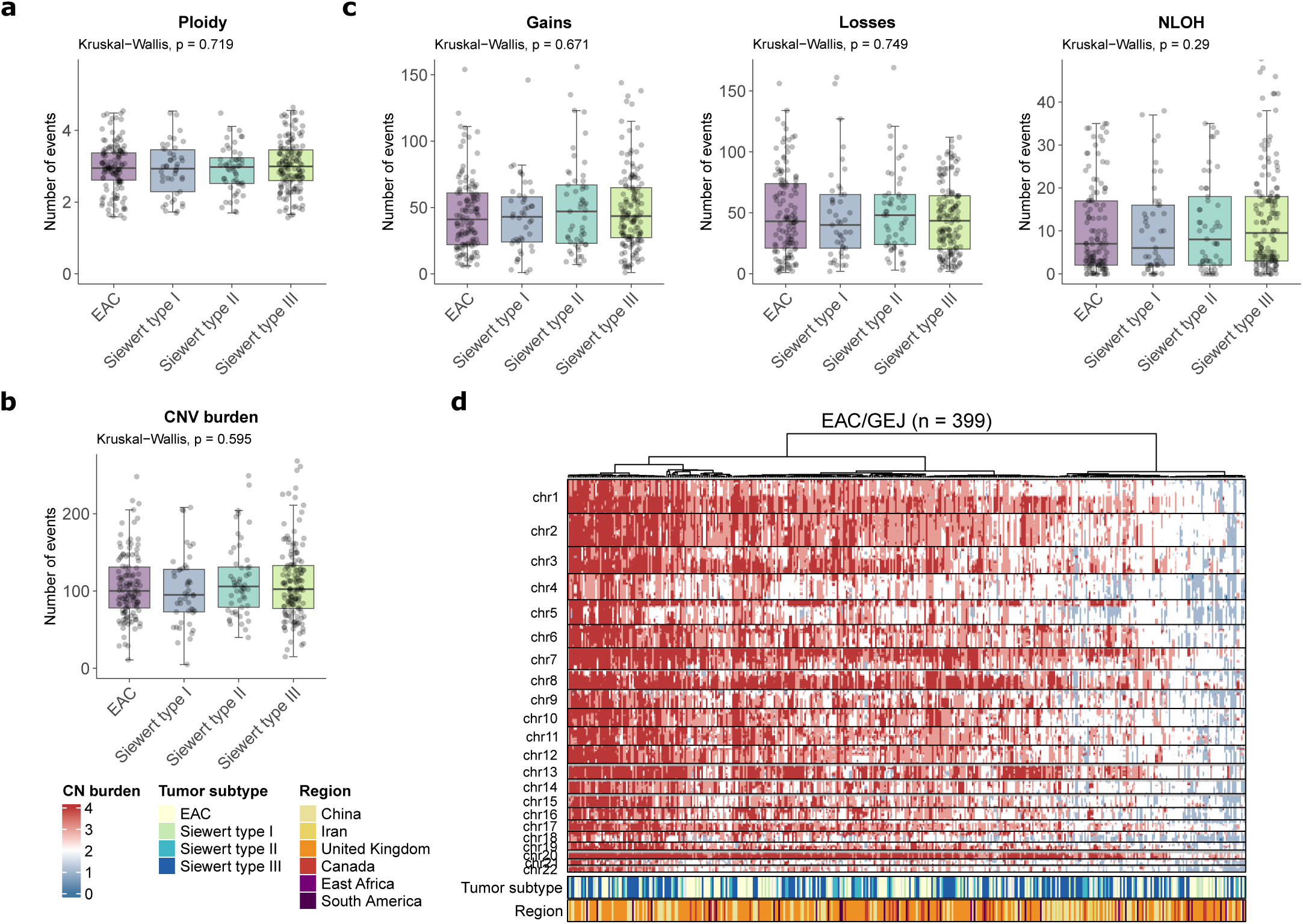
Copy number profile in EAC vs GEJ. **a-c,** Ploidy (**a**), copy number burden (**b**), and burden of gains, losses, and copy-neutral LOH (NLOH) (**c**) across EAC and GEJ Siewert types (n = 399 biologically independent samples). The Kruskal–Wallis test (two-sided) was used to test for global differences. Box-and-whisker plots are in the style of Tukey. The line within the box is plotted at the median, while upper and lower ends indicate 25th and 75th percentiles. Whiskers show 1.5 × IQR. The y axes were cut at 1.25× upper whisker for clarity. **d**, Copy number burden across the genomes of EAC/GEJ. Samples are sorted based on unsupervised hierarchical clustering of copy number counts, computed using Euclidean distance and Ward’s agglomerative procedure. The bottom panel indicates the tumor subtype and region per case.

## SUPPLEMENTARY TABLES

**Supplementary Table 1**. SBS, DBS, and ID burdens in EAC/GEJ samples.

**Supplementary Table 2**. Associations of copy number and mutation burdens with epidemiological and clinical features in EAC/GEJ tumors.

**Supplementary Table 3**. de novo SBS signatures extracted from EAC/GEJ cases.

**Supplementary Table 4**. de novo DBS signatures extracted from EAC/GEJ cases.

**Supplementary Table 5**. de novo ID signatures extracted from 486 EAC/GEJ cases.

**Supplementary Table 6**. Decomposition of de novo mutational signatures to COSMIC reference signatures.

**Supplementary Table 7**. Activities of de novo mutational signatures in EAC/GEJ cases.

**Supplementary Table 8**. Activities of decomposed signatures in EAC/GEJ cases.

**Supplementary Table 9**. Mutational signatures in early and late clonal mutations in EAC/GEJ tumors.

**Supplementary Table 10**. Associations of mutational signatures with epidemiological and clinical features in EAC/GEJ tumors (n=486).

**Supplementary Table 11**. Associations of mutational signatures with epidemiological and clinical features in EAC tumors (n=151).

**Supplementary Table 12**. Associations of mutational signatures with epidemiological and clinical features in GEJ tumors (n=335).

**Supplementary Table 13**. Associations of COSMIC mutational signatures with ancestry groups in the full cohort and by tumor subtype.

**Supplementary Table 14**. Detection of *Helicobacter pylori* reads in tumor DNA.

**Supplementary Table 15**. Associations of mutational signatures with age-standardized incidence rates of EAC/GEJ.

**Supplementary Table 16**. Fisher test results between EAC/GEJ drivers and epidemiological factors in the full dataset and across tumor subtypes.

**Supplementary Table 17**. Details of individual case collections.

**Supplementary Table 18**. Admixture analysis in EAC/GEJ samples.

**Supplementary Table 19**. Comparison of single base substitution signatures extracted by SigProfilerExtractor and mSigHdp.

**Supplementary Table 20**. Comparison of indel signatures extracted by SigProfilerExtractor and mSigHdp.

**Supplementary Table 21**. Associations of copy number clusters with epidemiological features.

