## Supplementary Material for "Convergent mutational processes in esophageal and junctional adenocarcinomas across eight countries"

**SUPPLEMENTARY INFORMATION INVENTORY**

This file contains the following:

Supplementary Note

Supplementary References

Supplementary Table Legends

Supplementary Figures

**SUPPLEMENTARY NOTE**

#### 1. Supplementary methods

##### 1.1. Extraction of de novo mutational signatures with SigProfilerExtractor

SBS Signatures were extracted in both SBS288 and SBS1536 contexts. These extend the original SBS96 contexts to capture distinct features, with SBS288 taking into account the transcribed, untranscribed, and intergenic regions of the genome and SBS1536 considering two bases on either side of the mutated one to form a pentonucleotide context. Signatures were extracted using both contexts, as it is not possible to predict *a priori* which classification will be best able to extract the particular combination of mutational signatures in any given landscape. This also provides additional confidence in the signatures extracted. The results from SBS288 and SBS1536 were very similar, with both extractions producing 12 signatures. Eleven signature pairs were highly similar with cosine similarities of 0.96 or greater (**Supplementary Figure 1**, **Supplementary Table 19**). The remaining pair (SBS1536G and SBS288E) had a lower cosine similarity, which is due to the SBS288 extraction failing to extract SBS103 as cleanly compared to the SBS1536 extraction.

##### 1.2. Extraction of de novo mutational signatures with mSigHdp

SBS96 and ID83 signatures were also extracted using mSigHdp to validate the SigProfilerExtractor results. For SBS96, mSigHdp extracted 15 signatures in total (**Supplementary Figure 2)** of which 12 signatures were similar to those extracted by SigProfilerExtractor (**Supplementary Table 19**) while three were unique to the mSigHdp extraction, Hdp.3, Hdp.11 and Hd.13.

Hdp.3 had a broad spectrum with many peaks in C>T, T>A, and T>C contexts. On default settings, it decomposed into COSMIC reference signatures SBS1, SBS5, SBS21, and SBS85. SBS21 is a signature associated with microsatellite instability, while SBS85 is the result of activation-induced cytidine deaminase (AID) in B-cells[1,2]. This result is unlikely given the tissue type and exclusion of MSI cases from the extraction. On optimised settings, it decomposed into COSMIC reference signatures SBS1, SBS5, SBS12, and SBS34, which is also unlikely, as SBS12 is thought to be an exogenous exposure found in Japanese kidney and liver cancers[3]. In a test attribution using SigProfilerAttribution, only 3 samples had low levels of SBS12 attributed, and no SBS34 was attributed at all. It is possible that Hdp.3 is instead an attempt to separate SBS111, as it has some similarity to the *de novo* versions SBS1536H, SBS288H, and Hdp.14 (0.85, 0.83 and 0.72 cosine similarity respectively), but lacking the C>A and

N[C>T]G sites. However, given that Hdp.3 did not have a feasible decomposition and could not be validated in either SigProfilerExtractor extraction, it was not accepted into the final panel of *de novo* signatures.

Hdp.11 decomposed to SBS1, SBS5, SBS8 and SBS17a/b with a cosine similarity of 0.93, and is likely the partial separation of the T>A peaks seen in SBS17a. As all signatures in the decomposition were part of the final signature panel used for attribution regardless, this additional mSigHdp signature has no impact on the results.

Hdp.13 was composed of C[C>T]G and A[C>T]G mutations, and on default settings decomposed into SBS87 (associated with Thiopurine chemotherapy treatment[4]) with a cosine similarity of 0.85. Review of the attribution showed that only one sample had a large number of mutations attributed to Hdp.13, and although extensive medical records were available for this individual, there was no evidence of any previous chemotherapy treatment. In addition, the C>G peaks of SBS87 where not present in Hdp.13. Taken together, this indicates that SBS87 is unlikely to be present in this cohort. When chemotherapy signatures were excluded from the decomposition, the signature decomposed into SBS1 and SBS5 with a cosine similarity of 0.82.

mSigHdp extracted 9 indel signatures in total (**Supplementary Figure 3)** compared to the five extracted by SigProfilerExtractor. Three mSigHdp were close matches with a corresponding SigProfilerExtractor (**Supplementary Table 20**). Of the remaining 6 signatures, Hdp.2 was another mixture of ID1 and ID2 with different proportions compared to the other ID1/ID2 containing signatures. Hdp6 and hdp8 represented the ID10 and ID9 components, which in SigProfiler are contained within ID83D. The last three unique signatures, hdp.5, hdp.9 and hdp.7, appear to be attempts to separate the component signatures of ID83C, which include ID2, ID4, ID8, ID9, and ID21. A combination of these three signature reconstituted ID83D with a cosine similarity of 0.95.

##### 1.3. Decomposition to reference signatures

The 12 *de novo* SBS1536 signatures were taken forward as the final panel of signatures used for decomposition to COSMIC reference signatures. Default decompositions of mutational signatures are often incorrect due to the presence of signatures that are not biologically feasible being included in the pool. SigProfilerAssignment includes parameters which can exclude specific groups of signatures that share an etiology, although care must be taken to only do so when relevant, for example, excluding MSI signatures when all such cases were removed prior to extraction. The optimised decomposition results were accepted for all signatures except SBS1536D. On default settings, this signature decomposes into SBS5, SBS17a, SBS17b, and SBS28. On optimised settings, SBS5 and SBS28 are replaced with SBS40b. SBS28 is excluded from the optimised decomposition as it is frequently found at high mutation burden in samples with signatures SBS10a and 10b, which are caused by mutations in polymerase epsilon[1]. More recently, it has been shown that SBS28 mutations can also be present in much lower burdens in POLE+ samples[5]. On the other hand, SBS40b is unlikely, as to date it has only been found in normal kidney tissue and clear cell renal cell carcinomas[3,6]. On this basis, the default decomposition was accepted for SBS1536D.

#### 2. Supplementary results

##### 2.1. Principal component analysis of mutation SBS96 counts and SBS signature attributions

Principal component analysis was conducted using the relative proportions of each of the SBS96 contexts. This shows no clustering on the basis of tumor type (**Supplementary Figure 4a),** which is consistent with other results that show that EAC and GEJ have a similar mutational landscape (**Figure 2**). Instead, the bulk of the variation can be explained by a higher proportion of T>C and T>G mutations (**Supplementary Figure 4b)** and a higher relative proportion of SBS17 and SBS17b attribution (**Supplementary Figure 4c-d)**.

##### 2.2. Convergent driver profiles among EAC and GEJ

Our analysis of 399 cases with available mutation and copy number information revealed 179 cancer genes harboring driver alterations in our dataset. Six genes presented alterations in >10% of cases, including *TP53* (81.7%), *CDKN2A* (19.6%), *KRAS* (13.8%), *SMAD4* (12%), *ARID1A* (10.7%), and *MYC* (10.3%) (**Figure 4**), which are commonly implicated in EAC[7,8] and gastric adenocarcinomas[9]. The driver profile was similar in EAC and GEJ, and across all clinical subtypes (**Supplementary Table 16**). Only minor differences were observed, including a higher frequency of *CDKN2A* mutations in EAC, whereas deletions in this gene were more prevalent in GEJ (**Figure 4c**).

Despite the high copy number instability and aneuploidy characteristic of these malignancies[7,10], no differences in copy number burdens and ploidy were found across EAC and GEJ (**Extended Data Figure 6**, **Supplementary Table 2**). Consistent with the homogeneous mutation profile across tumors, we found no association between specific lifestyle factors and the spectrum of driver mutations or copy number alterations compared to unexposed cases (**Supplementary Table 2,Supplementary Table *16***). These results further evidence the lack of a mutagenic effect of exposures such as tobacco and alcohol in EAC/GEJ. Despite regional variations in the mutational signature backgrounds, the landscape of driver alterations was overall uniform across geographical regions. Only *CCNE1* amplifications showed a significant enrichment in Chinese cases compared to other regions (**Figure 4d**).

##### 2.3. Copy number clustering analysis

EAC/GEJ are characterized by high copy number (CN) instability[10]. To assess the underlying patterns of CN variations, we performed unsupervised clustering analysis of the CN counts using Euclidean distance and Ward's agglomerative procedure (**Supplementary Figure 5**). The analysis revealed three main groups of samples. Cluster 1 (16% of EAC/GEJ cases, n = 65) was defined by polyploidy and high burden of genomic gains. Cluster 2 (57%, n = 226), the most prevalent group, was characterized by polyploidy and high burdens of genomic losses. Cluster 3 (27%, n = 108) presented near-diploid genomes, a lower overall CN burden, and high frequency of loss of heterozygosity. These clusters showed no significant association with tumor subtype, geographical region, or other epidemiological characteristics (**Supplementary Figure 5 Supplementary Table 21**), reinforcing the homogeneous genomic profiles between EAC and GEJ.

##### 2.4. Association between bile acid reflux signatures and obesity prior to diagnosis

We identified an association between obesity, assessed at the time of diagnosis, and reflux signatures SBS17a and SBS17b in EAC, but not in GEJ cases (**Figure 5, Supplementary Table *10*-Supplementary Table *12***). To determine whether the absence of this association in GEJ was confounded by cancer-induced weight loss, we evaluated historical obesity data available for a subset of patients. Specifically, we analyzed obesity status two years prior to diagnosis (n = 72) and at 40 years of age (n = 65) (**Supplementary Figure 6**). Despite the limited sample size, we observed a consistent trend toward higher SBS17a and SBS17b burdens in obese EAC cases in both time points, whereas no such trend was apparent in GEJ cases. These findings reinforce the link between obesity and bile acid reflux signatures in EAC, suggesting that our observations are not artifacts of disease-related weight loss.

##### 2.5. *Helicobacter pylori* detection and geographic heterogeneity

Given the proposed role of *Helicobacter pylori* infection in the etiology of GEJ, described specifically in East Asia[11,12], we evaluated the presence of *H. pylori* reads in tumor sequencing data (**Supplementary Table 14**). We identified 29 cases displaying *H. pylori* reads (**Supplementary Figure 7a**). Regression analysis revealed a significant enrichment of *H. pylori*-positive cases in China (18%, 14/107) and Iran (31%, 8/26) compared to other countries (2%, 7/348) (**Supplementary Figure 7b-c**). This provides molecular evidence of a potential role of the infection in cancer development.

It has been hypothesized that *H. pylori*-positive GEJ might constitute a distinct form of GEJ, resembling non-cardia gastric cancer more closely than EAC[11]. However, we found no evidence of distinct mutational patterns associated with *H. pylori* status, with no significant differences in signature distribution (**Supplementary Table 10-Supplementary Table *12***). These data suggest that, while *H. pylori* prevalence varies geographically and could hypothetically contribute to the local incidence of GEJ, the infection is not responsible for the distinct mutational patterns observed in China. Definitive assessment of a potential association between infection and mutational profiles would require data on historical *H. pylori* infections or eradication treatments, which were unavailable for this cohort.

#### SUPPLEMENTARY TABLE LEGENDS

**Supplementary Table 1.** SBS, DBS, and ID burdens in EAC/GEJ samples.

**Supplementary Table 2.** Associations of copy number and mutation burdens with epidemiological and clinical features in EAC/GEJ tumors.

**Supplementary Table 3.** de novo SBS signatures extracted from EAC/GEJ cases.

**Supplementary Table 4.** de novo DBS signatures extracted from EAC/GEJ cases.

**Supplementary Table 5.** de novo ID signatures extracted from 486 EAC/GEJ cases.

**Supplementary Table 6**. Decomposition of de novo mutational signatures to COSMIC reference signatures.

**Supplementary Table 7.** Activities of de novo mutational signatures in EAC/GEJ cases.

**Supplementary Table 8.** Activities of decomposed signatures in EAC/GEJ cases.

**Supplementary Table 9.** Mutational signatures in early and late clonal mutations in EAC/GEJ tumors.

**Supplementary Table 10.** Associations of mutational signatures with epidemiological and clinical features in EAC/GEJ tumors (n=486).

**Supplementary Table 11.** Associations of mutational signatures with epidemiological and clinical features in EAC tumors (n=151).

**Supplementary Table 12.** Associations of mutational signatures with epidemiological and clinical features in GEJ tumors (n=335).

**Supplementary Table 13.** Associations of COSMIC mutational signatures with ancestry groups in the full cohort and by tumor subtype.

**Supplementary Table 14.** Detection of *Helicobacter pylori* reads in tumor DNA.

**Supplementary Table 15.** Associations of mutational signatures with age-standardized incidence rates of EAC/GEJ.

**Supplementary Table 16.** Fisher test results between EAC/GEJ drivers and epidemiological factors in the full dataset and across tumor subtypes.

**Supplementary Table 17.** Details of individual case collections.

**Supplementary Table 18.** Admixture analysis in EAC/GEJ samples.

**Supplementary Table 19.** Comparison of single base substitution signatures extracted by SigProfilerExtractor and mSigHdp.

**Supplementary Table 20.** Comparison of indel signatures extracted by SigProfilerExtractor and mSigHdp.

**Supplementary Table 21.** Associations of copy number clusters with epidemiological features.

#### SUPPLEMENTARY FIGURES

**Supplementary Figure 1. Single-base substitution signatures extracted by SigProfilerExtractor.** All 12 singl-base substitution (SBS) de novo signatures extracted in SBS288 and SBS1536 format, shown side by side for comparison.

**Supplementary Figure 2. Single base substitution signatures extracted by mSigHdp.** All single-base substitution (SBS) de novo signatures extracted by mSigHDP.

**Supplementary Figure 3. Small insertion and deletion signatures extracted by mSigHdp.** All small insertion and deletion (ID) de novo signatures extracted by mSigHDP.

**Supplementary Figure 4. Principal component analysis.** Principal component analysis of EAC/GEJ cohort (n=486) performed on relative SBS96 mutation counts, colored by **a**, tumor subtype, **b**, relative proportion of SBS6 contexts, **c**, relative attribution of SBS17a, and **d**, relative attribution of SBS17b.

**Supplementary Figure 5. Copy number profile of EAC/GEJ clusters. a,** Unsupervised hierarchical clustering analysis of copy number counts in the EAC/GEJ cohort (n=399) using Euclidean distance and Ward's agglomerative procedure, displaying three clusters. **b,** Genome-wide segments showing major and minor allele counts in 15 randomly picked samples per copy number cluster. **c,** Frequencies of copy number clusters across tumor subtypes and geographical regions. The number of positive and negative samples is indicated. Significance was assessed using Fisher's exact test. **d**, Ploidy, copy number burden and burden of gains, losses and copy-neutral LOH (NLOH) across clusters. The Kruskal–Wallis test (two-sided) was used to test for global differences. Box-and-whisker plots are in the style of Tukey. The line within the box is plotted at the median, while upper and lower ends indicate 25th and 75th percentiles. Whiskers show 1.5 × IQR.

**Supplementary Figure 6. Associations between obesity prior to diagnosis and bile acid signatures.** Mutational burdens for SBS17a and SBS17b signatures in cases reporting obesity prior to diagnosis in EAC (**a, b**) and GEJ (**c, d**) tumors. Variables employed include obesity two years prior to diagnosis (**a**, **c,** n = 72 independent samples) and at 40 years of age (**b**, **d**, n = 65 independent samples). The Kruskal–Wallis test (two-sided) was used to test for global differences. Box-and-whisker plots are in the style of Tukey. The line within the box is plotted at the median, while the upper and lower ends indicate the 25th and 75th percentiles. Whiskers show 1.5× IQR. The y axes were cut at 1.25× upper whisker for clarity. Bar plots indicate the frequencies of dichotomized signatures.

**Supplementary Figure 7. *Helicobacter pylori* detection in tumor DNA. a**, Species counts for Helicobacter pylori obtained from Metaphlan and Bracken algorithms. Samples with detected *H. pylori* reads from both methods are considered positive. The number of samples positive for each algorithm is displayed at the right. **b**, Frequencies of samples positive for *H. pylori* detection across geographical regions. The number of positive and negative samples is indicated. Significance was assessed using Fisher's exact test. **c**, Associations between H. pylori positivity and geographical region in EAC/GEJ, measured by logistic regression analysis. Dots filled in color indicate significant associations (adjusted p-value $< 0.05$). Regressions were corrected for age of diagnosis, sex, tumor subtype, and tumor purity.

### Supplementary Figure 1

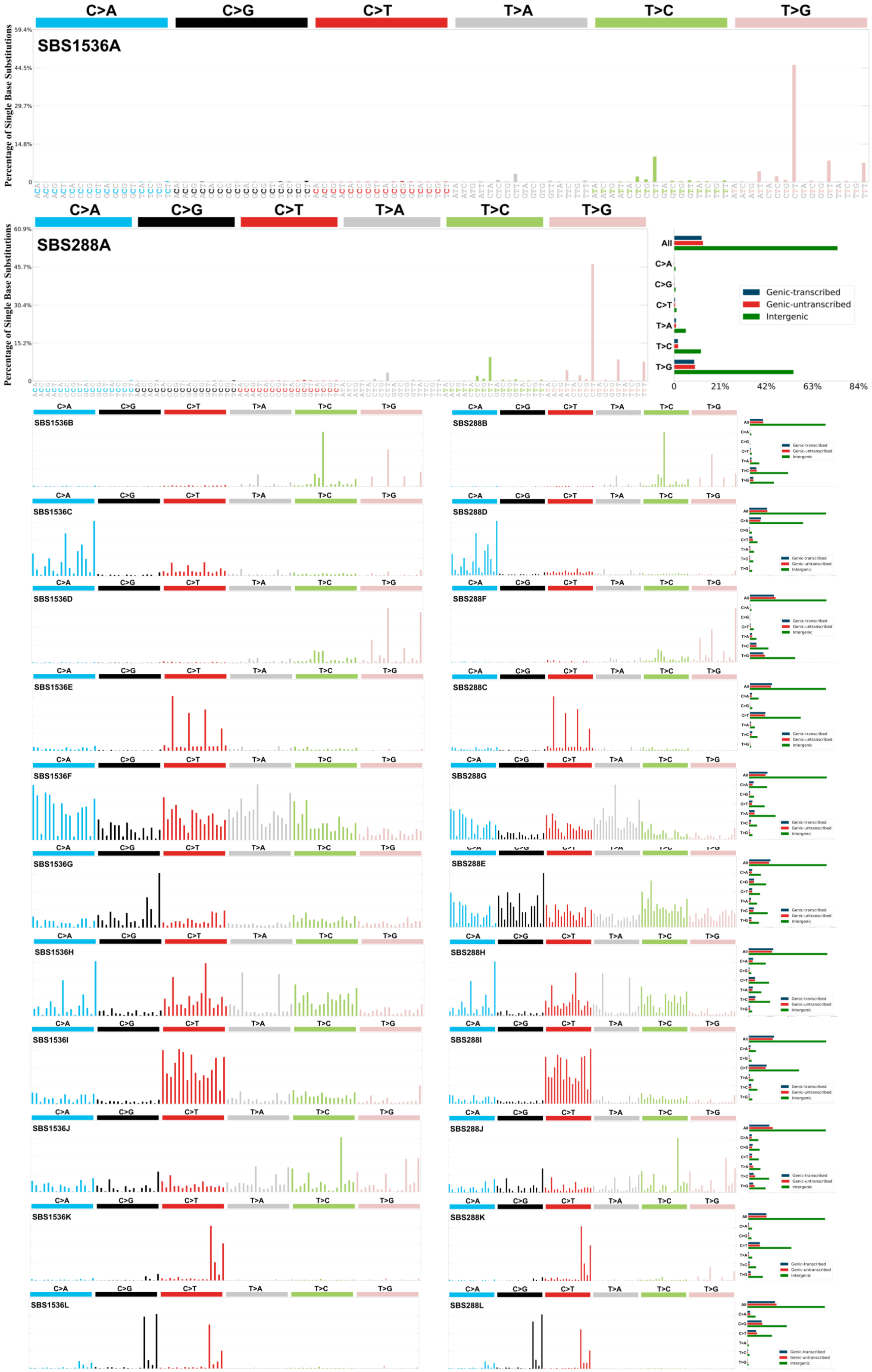

### Supplementary Figure 2

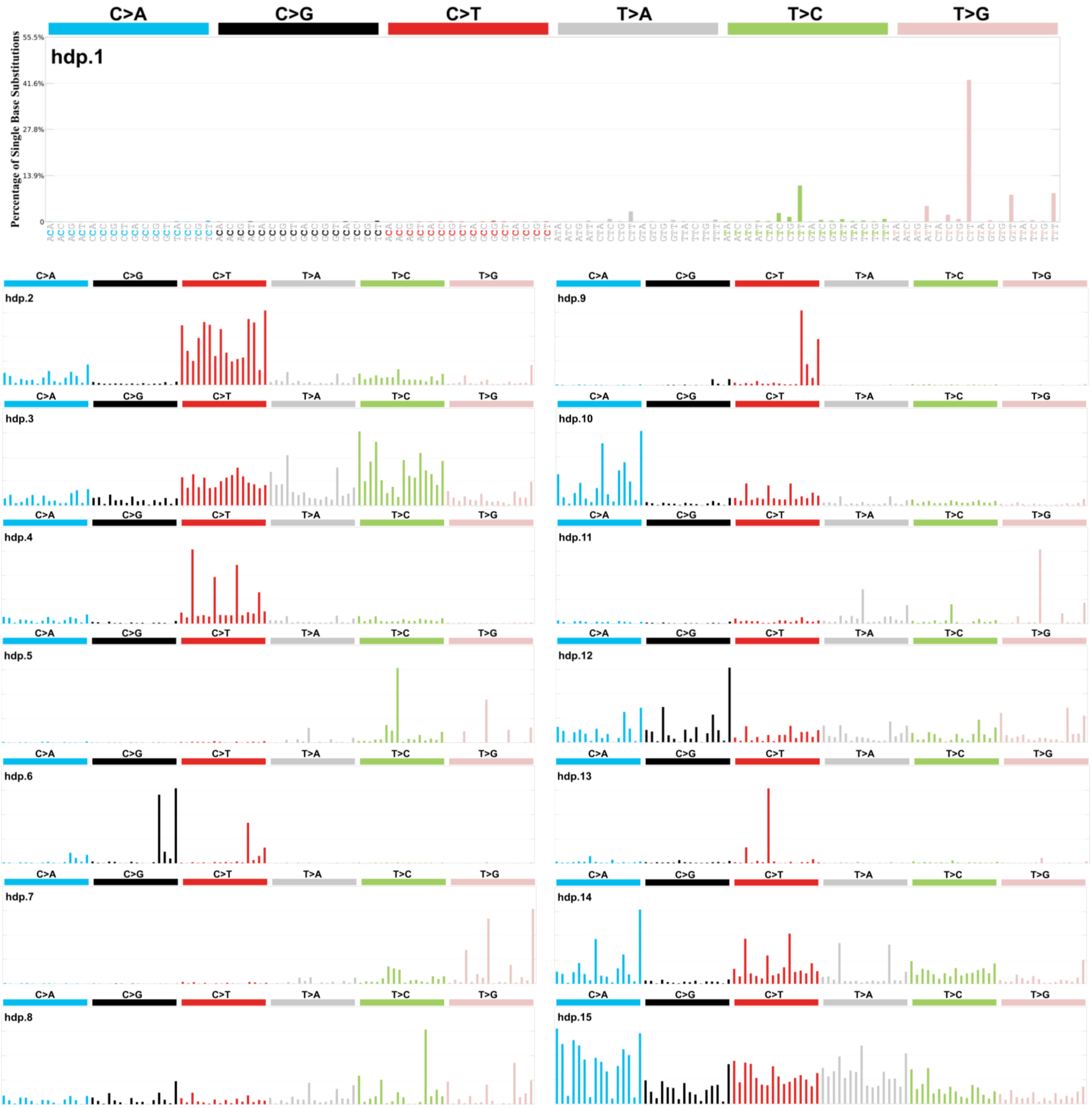

### Supplementary Figure 3

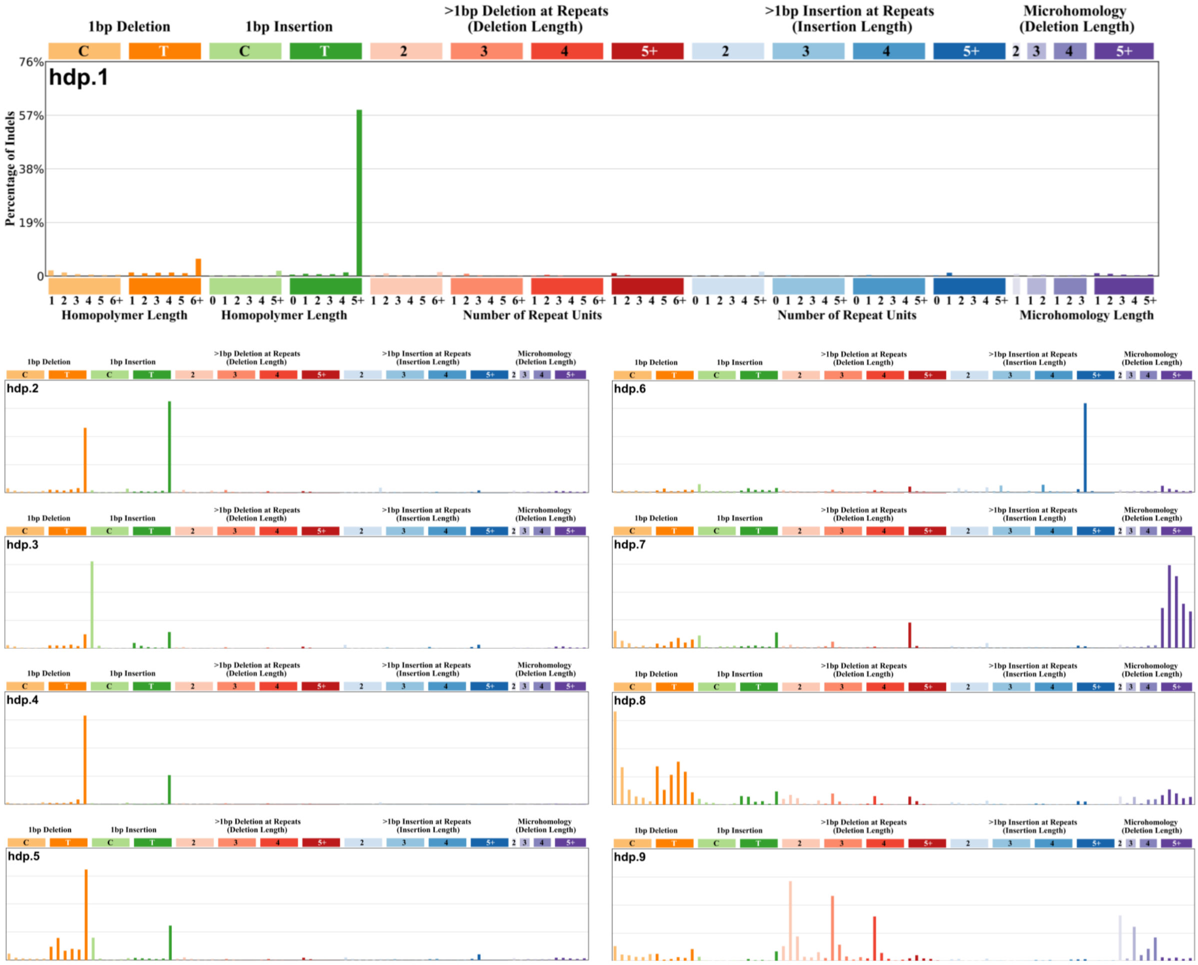

### Supplementary Figure 4

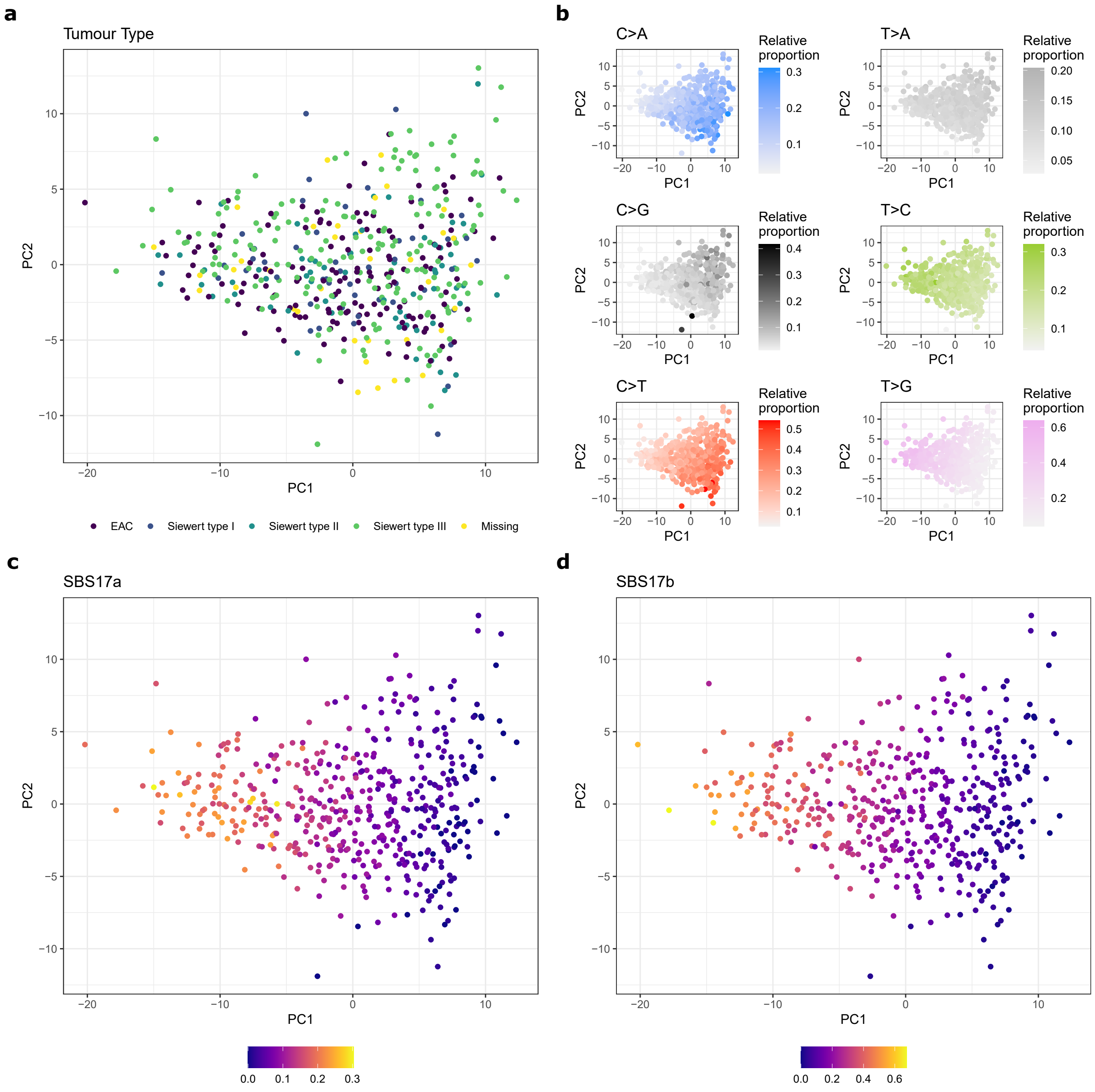

### Supplementary Figure 5

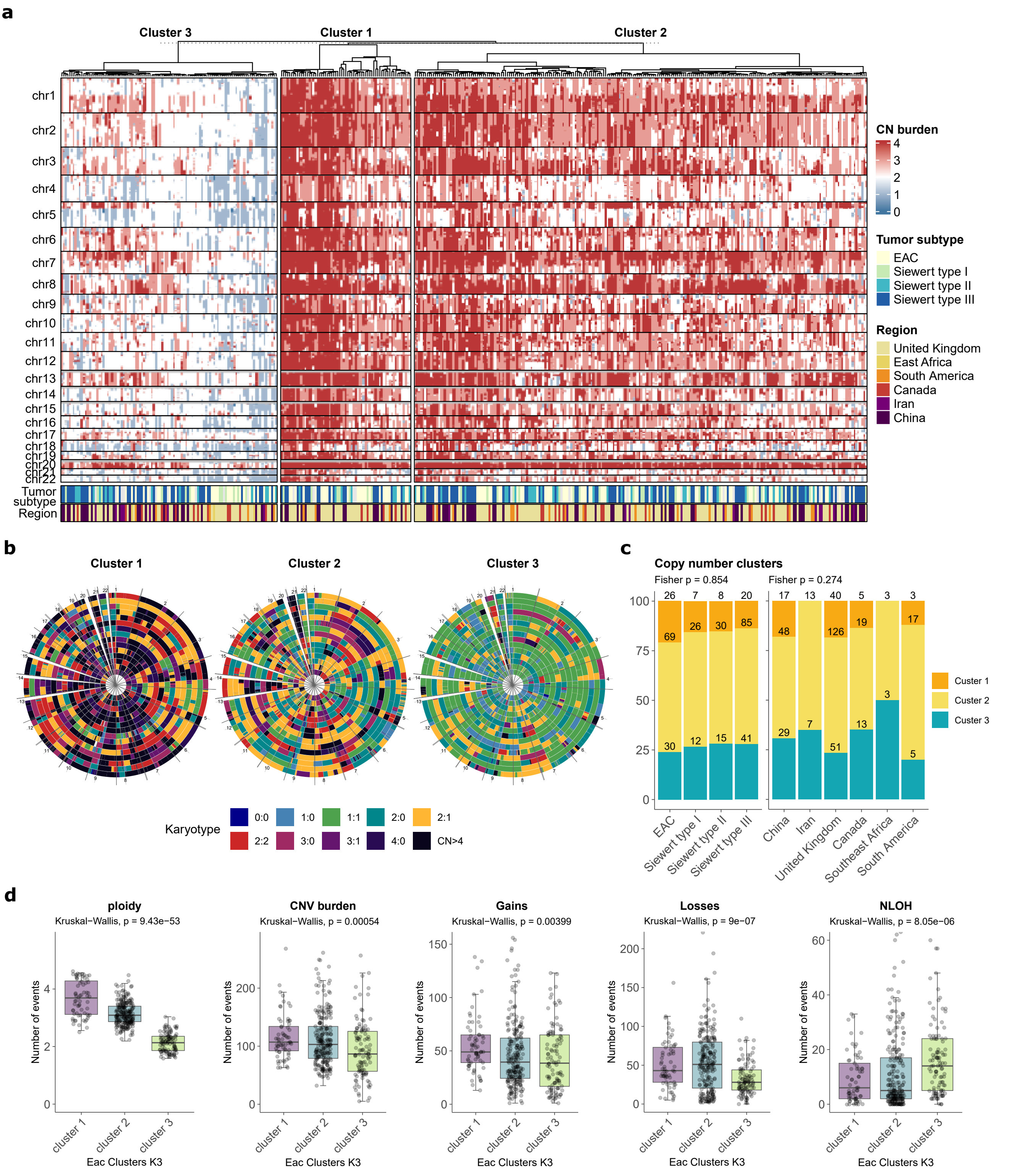

#### Supplementary Figure 6

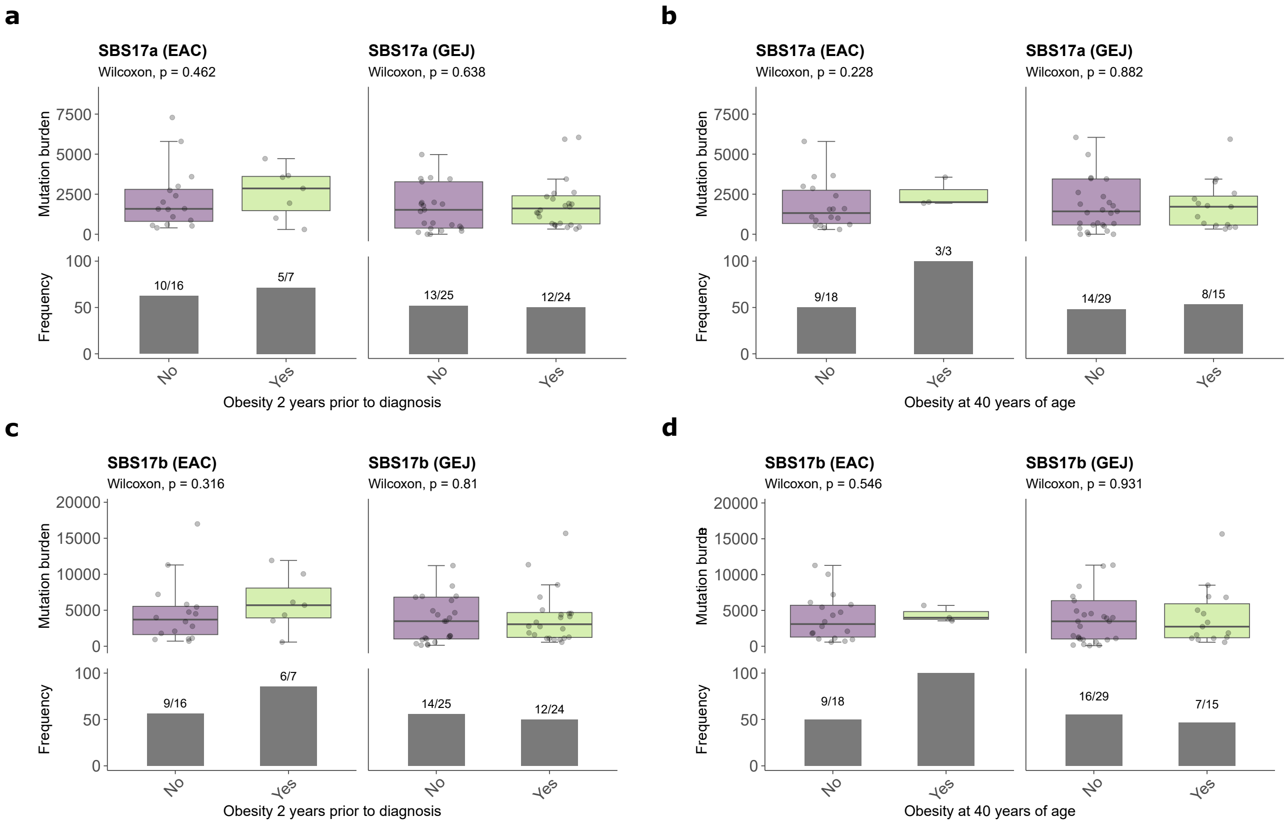

### Supplementary Figure 7

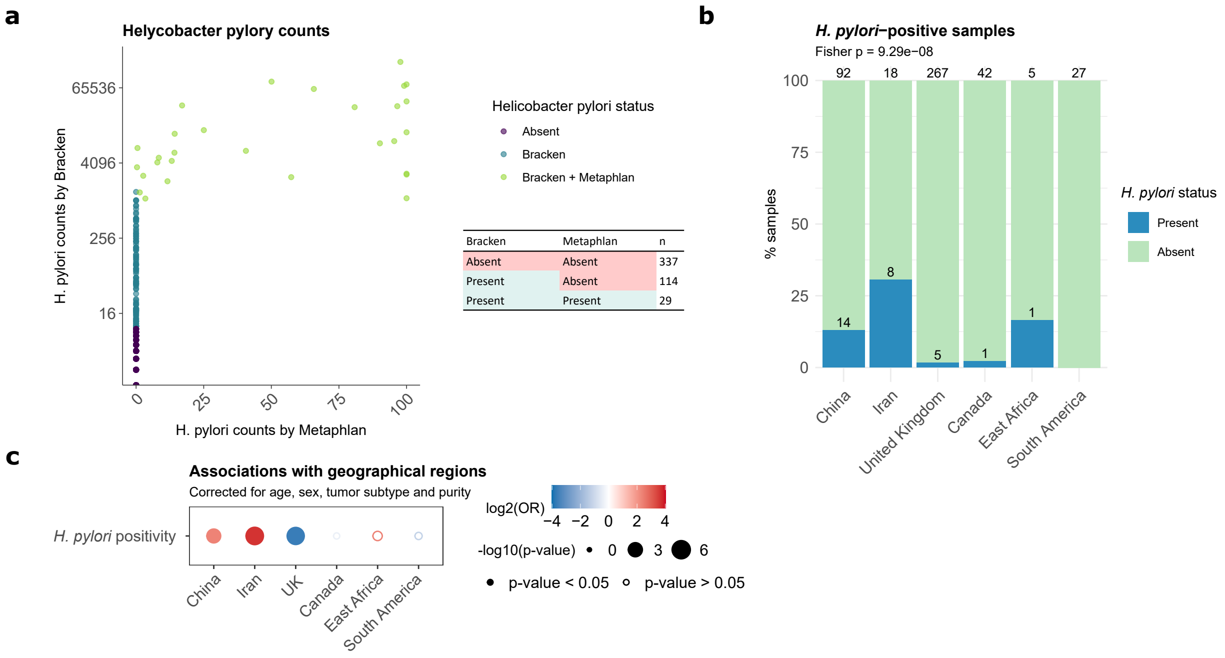
